# Prediction of pharmacist medication interventions using medication regimen complexity

**DOI:** 10.1101/2024.10.23.24316001

**Authors:** Bokai Zhao, Ye Shen, John W. Devlin, David J. Murphy, Susan E. Smith, Brian Murray, Sandra Rowe, Andrea Sikora

**Affiliations:** University of Georgia College of Public Health, Epidemiology & Biostatistics, Athens, GA, USA; University of Georgia, College of Public Health, Epidemiology & Biostatistics, Athens, GA, USA; Northeastern University School of Pharmacy, Boston, MA; Brigham and Women’s Hospital, Division of Pulmonary and Critical Care Medicine, Boston, MA; Emory University, Division of Pulmonary, Allergy, Critical Care and Sleep Medicine, Atlanta, GA, USA; University of Georgia College of Pharmacy, Department of Clinical and Administrative Pharmacy, Athens, GA, USA; University of Colorado Skaggs School of Pharmacy, Aurora, CO, USA; Oregon Health Sciences University, Portland, OR, USA

**Keywords:** Medication regimen complexity, medication safety, critical care, pharmacist

## Abstract

**Background:** Critically ill patients are managed with complex medication regimens that require medication management to optimize safety and efficacy. When performed by a critical care pharmacist (CCP), discrete medication management activities are termed medication interventions. The ability to define CCP workflow and intervention timeliness depends on the ability to predict the medication management needs of individual intensive care unit (ICU) patients. The purpose of this study was to develop prediction models for the number and intensity of medication interventions in critically ill patients.

**Methods:** This was a retrospective, observational cohort study of adult patients admitted to an ICU between June 1, 2020 and June 7, 2023. Models to predict number of pharmacist interventions using both patient and medication related predictor variables collected at either baseline or in the first 24 hours of ICU stay were created. Both regression and supervised machine learning models (Random Forest, Support Vector Machine, XGBoost) were developed. Root mean square derivation (RMSE), mean absolute error (MAE), and symmetric mean absolute percentage error (sMAPE) were calculated.

**Results:** In a cohort of 13,373 patients, the average number of interventions was 4.7 (standard deviation (SD) 7.1) and intervention intensity was 24.0 (40.3). Among the ML models, the Random Forest model had the lowest RMSE (9.26) while Support Vector Machine had the lowest MAE (4.71). All machine learning models performed similarly to the stepwise logistic regression model, and these performed better than a base model combining severity of illness with medication regimen complexity scores.

**Conclusions:** Intervention quantity can be predicted using patient-specific factors. While inter-institutional variation in intervention documentation precludes external validation, our results provide a framework workload modeling at any institution.

## Introduction

Medication therapy in the intensive care unit (ICU) is notoriously complex and high-risk requiring nuanced oversight and management.^1–6^ It has been shown that one in six ICU medication errors require intervention by a critical care pharmacist (CCP).^7^ Tailored medication interventions in the context of comprehensive medication management (CMM) performed by CCPs reduce preventable adverse drug events (ADEs) by 70%.^8–11^ However, staffing constraints and shortages combined with unpredictable and sometimes surging patient volumes may result in ICU patients receiving inconsistent CCP care.^12, 13^ Thus, predicting and prioritizing those patients with the highest need for CCP medication intervention(s) has important ramifications for workload justification and redesign at the administrative level and patient triage at the clinician level.^1^

The medication regimen complexity-intensive care unit (MRC-ICU) score was designed to quantify the complexity of a critically ill patient’s medication regimen in a way that reflects the cognitive services provided by CCPs. This score has been related to both patient-centered outcomes, including mortality,^14^ length of stay^15^, ICU complications (e.g., fluid overload^16–18^), duration of mechanical ventilation, and drug-drug interactions, and pharmacist workload, including intervention quantity and intensity. ^19–25^ Moreover, application of machine learning approaches in the context of MRC-ICU have revealed new insights in how medications relate to patient outcomes.^26, 27^ Finally, this score showed a stronger relationship to medication interventions as compared to traditional severity of illness indicators.^28^

However, the predictive capability of medication complexity scores like MRC-ICU for CCP interventions that considers mediating factors like patient severity of illness has never been evaluated. The purpose of this study was to develop prediction methods for total medication interventions during a patient’s ICU stay. We hypothesized that higher MRC-ICU scores and higher severity of illness would be predictive for greater CCP medication interventions.

## Methods

### Study Population

This was a retrospective, observational study that was reviewed by the University of Georgia (UGA) Institutional Review Board (IRB) and determined to be exempt from IRB oversight (Project00001541). All methods were performed in accordance with the ethical standards of the UGA IRB and the Helsinki Declaration of 1975. Patient data were obtained via the Oregon Clinical and Translational Research Institute, which houses Epic^→^ electronic health record (EHR) data from Oregon Health and Science University (OHSU) Hospital. OHSU is a 576 bed academic medical center with cardiothoracic, trauma/surgery, neurocritical care, and medical intensive care units. Pharmacists are integrated into the medical teams to provide CMM, including presence on multiprofessional rounds. A total of 13,373 patients aged 18 years or older were identified between June 1, 2020 and June 7, 2023. Data from the first ICU admission per each patient were included. Patients were excluded if it was not their index ICU admission, the ICU stay was less than 24 hours, or if the patient was placed on comfort care within the first 24 hours of their ICU stay.

### Variables

The primary outcome was total medication interventions logged in Epic^→^ as i-Vents by a critical care pharmacist (CCP) during the patient’s ICU stay. The EHR was queried for relevant patient demographic information, medication information, and patient outcomes. Baseline characteristics including age, sex, race, ICU type, admission diagnosis, APACHE II score at 24 hours, and SOFA score at 24 hours, 48 hours, and 72 hours were collected. The MRC-ICU score was calculated at 24 hours, 48 hours, and 72 hours of the ICU stay. The MRC-ICU consists of 35 discrete medication categories with each category assigned a weighted value and then summed to create a score for a patient’s regimen at the given time point.^29^ For a patient prescribed vancomycin, norepinephrine, and docusate, they would be given 3 points for vancomycin, 1 point for norepinephrine, and 1 point for docusate for a total score of 5.

Interventions are logged by CCPs as part of routine care. The general expectation is that CCPs log interventions performed and categorize them using a standard categorization system that consists of 49 distinct categories.^20^ These categories are provided in **Supplemental Table 1**.

Additionally, these interventions were categorized into low, moderate, and high intensity intervention categories using a previously validated rating system to calculate intervention intensity.^13^ The composite score was equal to: (the number of low-intensity interventions) + 5*(the number of moderate-intensity interventions) + 25*(the number of high-intensity interventions). The weights were chosen for the three intensities of intervention to minimize an overlap of scores for different compositions of number of interventions.^13^

Following a literature review of pharmacist interventions in the ICU, potential predictor variables were identified by investigator consensus to include in each regression model. These variables included the following: baseline characteristics (age, sex, body mass index, ICU type, and admission diagnosis category) and 24-hour variables (sequential organ failure assessment (SOFA) at 24 hours, Acute Physiology and Chronic Health Evaluation II (APACHE II) at 24 hours, MRC-ICU at 24 hours, presence of delirium as indicated by positive confusion assessment method for the ICU (CAM-ICU) score.

### Analysis

Due to the hypothesis-generating nature of this study, no attempt was made to estimate the sample size. All eligible patients from the available database were included to maximize statistical power of the predictive models developed. Descriptive statistics were performed for relevant variables. Continuous variables were summarized by the mean and standard deviation, and categorical variables reported count and proportion of the total population. Clinical characteristics between patients with high and low interventions (defined as ≤ 5 (median) or not) were compared using either Student’s t-test or Chi-square test, as appropriate. Univariate analysis was performed on baseline variables to detect potential important variables for predicting intervention quantity and intensity. A two-sided p-value less than 0.05 was used to determine statistical significance for all outcomes. All analyses were performed using *R* (version 4.1.2).

A histogram for intervention count was plotted (see **Supplemental Figure 1**). MRC-ICU score was plotted by ICU day (see **Supplemental Figure 2** and **3**). High and low interventions were differentiated based on median and visual analysis. Mortality and intervention total were plotted by MRC-ICU decile (see **Supplemental Figure 4** and **5**, respectively). A composite score of severity of illness (as described by SOFA or APACHE II) and medication regimen complexity (as described by MRC-ICU) was plotted against interventions (see **Supplemental Figure 6**).

A total of six different prediction models were developed to predict the primary outcome (number of interventions during hospital stay) and secondary outcome (intensity score for interventions during hospital stay). These models included traditional regression (negative binomial regression with full predictors, stepwise selected predictors, and a simple model of two key predictors (MRC-ICU and SOFA score only)), and three machine learning models including Random Forest, Support Vector Machine (SVM), and XGBoost. Collinearity assessment was conducted via the variance inflation factor (VIF) or correlation matrix (see **Supplemental Table 2**).

Multiple imputation with 10 imputations per variable was applied for all missing data. Each model was trained on 10 imputed training datasets, and predictions were made on the corresponding 10 imputed testing datasets. The results from the 10 sub-models were pooled together based on Rubin’s rule. Univariate and multivariate analysis on full variables were performed on the 10 different imputed datasets. For each model, 80% of the observations were used as training data, and 20% were reserved as testing data.

### Traditional regression models

Multivariable generalized linear models (GLM) were developed to evaluate the relationship of MRC-ICU on ICU day 1 with total interventions. Both GLM with Poisson distribution and negative binomial distribution were explored (see **Supplemental Appendix A**). For variable selection, to ensure a narrower and more precise selection, model selection using Bayesian information criterion (BIC) with the additional option k=log(n) in a stepwise algorithm for each imputed dataset was performed. In the case of different variables being selected across the imputed datasets, the frequency of each variable being selected was recorded followed by the Wald test to determine whether each variable should remain in the final model in a stepwise process. A temporal analysis was conducted for changes in SOFA and MRC-ICU score (see **Appendix B**).

### Supervised machine learning models

During the model training for Random Forest, SVM and XGBoost, 5-fold cross validation was used to select hyperparameters. With those optimal hyperparameters, the trained model was fitted again on the whole training set. For Random Forest, number of trees and number of variables randomly sampled as candidates at each split were tuned. For SVM, linear kernel and cost of constraints violation were tuned. For XGBoost, maximum depth of a tree and maximum number of boosting iterations were tuned. Feature importance plots for Random Forest, SVM and XGBoost were plotted. Because ten different models were used on each imputed dataset, ten different feature importance lists were generated for each.

### Model evaluation

RMSE, MAE, and sMAPE were calculated for each set of predictions, and the results were pooled together. To mitigate the influence of outliers and skewed error distributions on performance metrics, the 95% quantile Root Mean Squared Error (RMSE), mean absolute error (MAE), and symmetric mean absolute percentage error (sMAPE) were calculated to provide a more robust assessment. Rate ratios (RR=e^β^, β is the estimated coefficient from the regression model) for the simple and stepwise model were calculated after variable selection and were reported for predictors of interest along with 95% confidence intervals (CI). The 95% quantile for both RMSE and MAE refer to the values below which 95% of the error values fall (with the remaining 5% as extreme errors), with lower values indicating better performance.

SHAP (SHapley Additive exPlanations) is a method based on game theory for explaining individual predictions from machine learning models.^30^ SHAP assigns a value to each feature, representing its impact on the prediction when that feature is included in the model compared to its absence, and thus provides a means to understand the contribution of each feature to the model’s prediction for a particular instance. SHAP values were calculated for each individual prediction, and SHAP bee swarm charts were used to visualize the feature impact.^31, 32^ Given that the feature importance plots and associated predictions were highly uniform across imputed datasets, the bee swarm chart were plotted based on on-fold of imputed dataset with the lowest RMSE for concise interpretation.

### Subgroup Analysis and Error Distribution

A subgroup analysis was conducted by categorizing interventions into high, medium, or low groups to evaluate the performance of prediction models across different subgroups. This approach was chosen because despite similar overall performance metrics found for each model, the distribution of predicted values varied significantly. We hypothesized that model performance might vary across intervention groups (e.g., one model might perform well in the low intervention group but poorly in the high intervention group whereas another model might show the opposite performance trend, resulting in similar overall performance metrics despite differing subgroup performance). To explore these differences, we analyzed the error distribution for each model. For models with comparable overall Root Mean Squared Error (RMSE), we conducted detailed subgroup analyses and plotted the error distributions for each model. Additional details are available in **Appendix C**.

## Results

A total of 13,373 patients were included in the analysis. The average age was 59.8 ± 17.4 years with an APACHE II score of 9.1 ± 4.1 at 24 hours and MRC-ICU score of 5.3 ± 3.9 at 24 hours. The overall mortality rate was 10.2% with 55.8% undergoing mechanical ventilation at 24 hours. Population characteristics are summarized in **Table 1**. The mean number of pharmacist interventions during the ICU stay was 4.7 ± 7.1 (see **Table 2**). Additionally, **Supplemental Table 3** provides a characterization of medication interventions by type and intensity level with discontinuation of clinically unwarranted medications (13.4%) and dosage adjustments (12.1%) being the top two categories.

**Table 1.** Population characteristics and outcomes.

|  | Total Cohort,<br>n=13,373 | High Interventions,<br>n=5,757 | Low Interventions,<br>n=7,616 | p-value |
| --- | --- | --- | --- | --- |
| Admission Demographics |  |  |  |  |
| Age | 59.8 (17.4) | 58.5 (16.7) | 60.8 (17.9) | <0.001 |
| Sex - Male | 7965 (59.6) | 3498 (60.8) | 4467 (58.7) | 0.01 |
| BMI | 29.4 (5.4) | 29.4 (8.5) | 29.5 (7.3) | 0.86 |
| ICU Type |  |  |  |  |
| Cardiac | 4458 (33.3) | 1914 (33.2) | 2544 (33.4) | <0.001 |
| Surgical/Trauma | 3807 (28.4) | 1686 (29.2) | 2121 (27.8) |  |
| Neurosciences | 2915 (21.8) | 1122 (19.4) | 1793 (23.5) |  |
| Medical | 2193 (16.4) | 1035 (17.9) | 1158 (15.2) |  |
| Admission Diagnosis |  |  |  |  |
| Cardiovascular | 2897 (21.6) | 1134 (19.7) | 1763 (23.1) | <0.001 |
| Gastrointestinal | 717 (5.3) | 348 (6.0) | 369 (4.8) |  |
| Hematologic | 277 (2.0) | 145 (2.5) | 132 (1.7) |  |
| Hepatic | 228 (1.7) | 148 (2.5) | 80 (1.0) |  |
| Infection | 616 (4.6) | 445 (7.7) | 171 (2.2) |  |
| Neoplasm | 772 (5.7) | 285 (4.9) | 487 (6.3) |  |
| Respiratory | 204 (1.5) | 65 (1.1) | 139 (1.8) |  |
| Sepsis | 377 (2.8) | 215 (3.7) | 162 (2.1) |  |
| Shock | 227 (1.7) | 153 (2.6) | 74 (0.9) |  |
| Trauma | 843 (6.3) | 327 (5.6) | 516 (6.7) |  |
| Other | 6215 (46.4) | 1635 (28.4) | 3723 (48.8) |  |
| 24 hour variables |  |  |  |  |
| MRC-ICU at 24 hours | 5.3 (3.9) | 7.0 (4.3) | 4.1 (3.0) | <0.001 |
| MRC-ICU at 48 hours | 5.0 (3.9) | 6.9 (4.3) | 3.5 (2.7) | <0.001 |
| MRC-ICU at 72 hours | 4.9 (3.8) | 6.6 (4.2) | 3.3 (2.5) | <0.001 |
| SOFA at 24 hours | 3.3 (3.0) | 4.3 (3.1) | 2.5 (2.7) | <0.001 |
| SOFA at 48 hours | 2.3 (2.8) | 3.4 (3.1) | 1.5 (2.2) | <0.001 |
| SOFA at 72 hours | 2.0 (2.6) | 2.9 (2.9) | 1.2 (1.9) | <0.001 |
| APACHE II at 24 hours | 9.1 (4.1) | 10.6 (4.3) | 7.9 (3.5) | <0.001 |
| Invasive mechanical ventilation in 24 hours | 7469 (55.8) | 4320 (75.0) | 3149 (41.3) | <0.001 |
| CAM-ICU in 24 hours | 10907 (81.5) | 5250 (91.1) | 5657 (74.2) | <0.001 |
| CRRT in 24 hours | 659 (4.9) | 573 (9.9) | 86 (1.1) | <0.001 |
| Outcomes |  |  |  |  |
| Hospital mortality | 1376 (10.2) | 878 (15.2) | 498 (6.5) | <0.001 |
| **Hospital length of stay (days)** | 12.79 (15.8) | 12.74 (15.5) | 12.82 (16.1) | 0.77 |
| **ICU length of stay (days)** | 4.6 (5.8) | 7.0 (7.8) | 2.8 (2.1) | <0.001 |
| Data are presented as n (%) or mean (standard deviation) unless otherwise designated. |  |  |  |  |
| SOFA: sequential organ failure assessment, APACHE II: Acute Physiology and Chronic Health Evaluation; ICU: intensive care unit |  |  |  |  |
| High interventions were defined as $\geq 5$ (median) and low as $< 5$. | | | | |

**Table 2.** CCP intervention data.

|  | Total Cohort,<br>n=13,373 | High Interventions,<br>n=5,757 | Low Interventions,<br>n=7,616 | p-value |
| --- | --- | --- | --- | --- |
| *CCP intervention data* |  |  |  |  |
| **Total interventions per ICU stay** | 4.7 (7.1) | 9.0 (9.0) | 1.5 (1.4) | <0.001 |
| **Total interventions per hospital stay** | 7.4 (9.6) | 14.0 (11.7) | 2.4 (1.6) | <0.001 |
| **Day 1 interventions** | 2.8 (2.3) | 3.7 (2.8) | 1.7 (0.9) | <0.001 |
| **Day 2 interventions** | 2.0 (1.7) | 2.4 (2.0) | 1.3 (0.6) | <0.001 |
| **Day 3 interventions** | 2.1 (1.7) | 2.3 (1.8) | 1.2 (0.5) | <0.001 |
| **Day 4 interventions** | 2.0 (1.6) | 2.1 (1.7) | 1.2 (0.5) | <0.001 |
| **Day 5 interventions** | 2.0 (1.6) | 2.1 (1.6) | 1.2 (0.5) | <0.001 |
| **Day 6 interventions** | 2.0 (1.6) | 2.1 (1.7) | 1.1 (0.4) | <0.001 |
| **Day 7 interventions** | 2.0 (1.7) | 2.0 (1.7) | 1.2 (0.5) | <0.001 |
| **Intervention intensity over ICU stay** | 24.0 (40.3) | 45.2 (52.7) | 8.1 (12.7) | <0.001 |
| **Day 1 intervention intensity** | 15.5 (20.4) | 20.8 (24.1) | 9.8 (12.9) | <0.001 |
| **Day 2 intervention intensity** | 10.0 (15.1) | 11.8 (17.0) | 6.4 (9.2) | <0.001 |
| **Day 3 intervention intensity** | 10.4 (15.2) | 11.7 (16.5) | 6.1 (8.4) | <0.001 |
| **Day 4 intervention intensity** | 9.5 (13.9) | 10.4 (14.8) | 5.7 (8.1) | <0.001 |
| **Day 5 intervention intensity** | 9.1 (12.5) | 9.5 (13.0) | 5.7 (7.8) | <0.001 |
| **Day 6 intervention intensity** | 9.4 (13.1) | 9.9 (13.5) | 4.7 (6.4) | <0.001 |
| **Day 7 intervention intensity** | 9.3 (13.4) | 9.6 (13.5) | 5.6 (11.2) | 0.01 |
| Data are presented as n (%) or mean (standard deviation) unless otherwise designated. |  |  |  |  |
| SOFA: sequential organ failure assessment, APACHE II: Acute Physiology and Chronic Health Evaluation; ICU: intensive care unit |  |  |  |  |
| High interventions were defined as $\geq 5$ (median) and low as $< 5$. | | | | |

Following multivariable analysis, it was observed that for every one point increase in the MRC-ICU, SOFA, and APACHE II score, there was approximately 10% more interventions made for the patient (see **Table 3**). **Supplemental Table 4** reports the final regression model for prediction of total interventions with a total of 12 variables, and **Supplemental Table 5** provides the model for a simple model of SOFA and MRC-ICU.

**Table 3.**
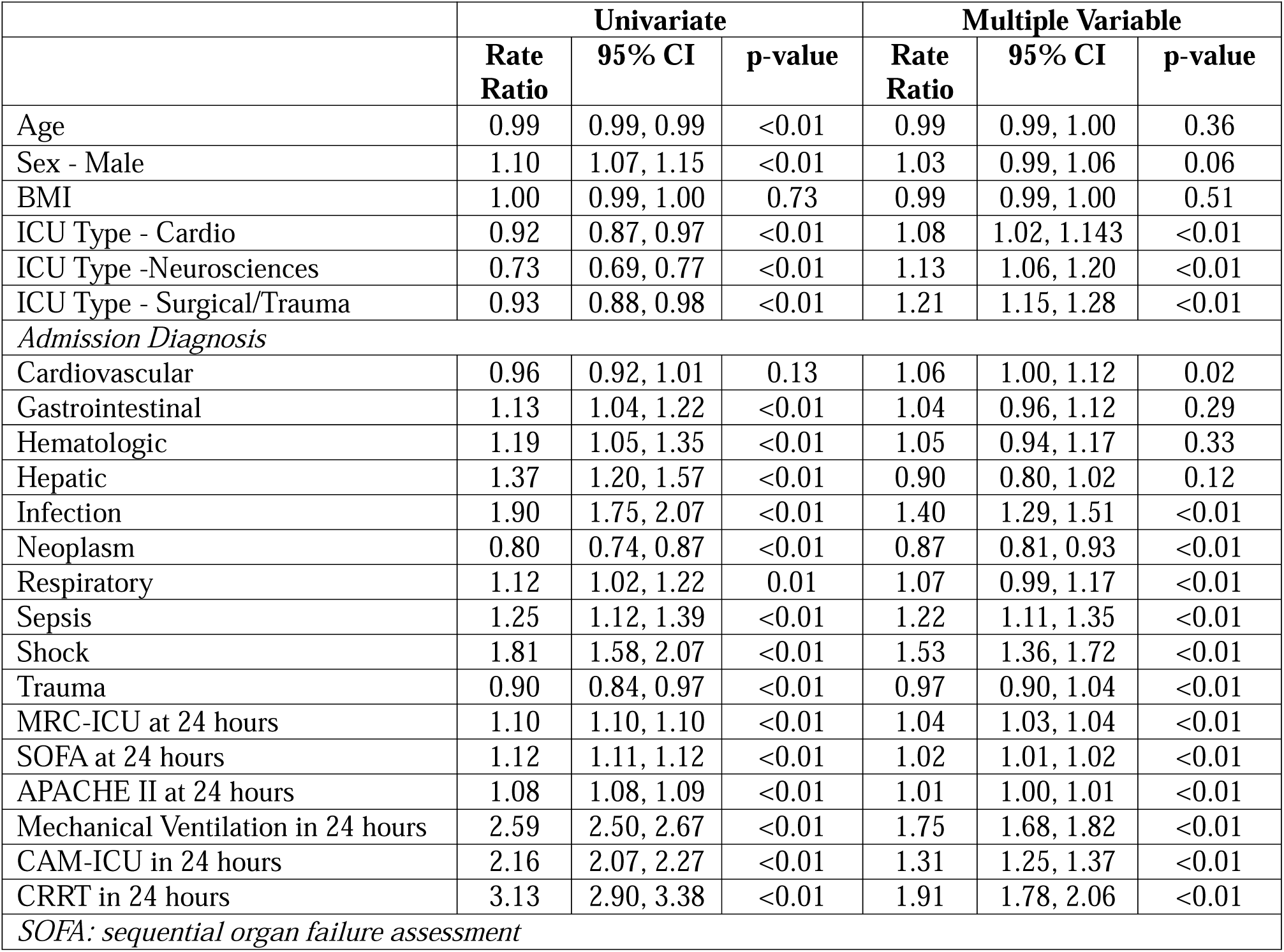
Univariate and multivariable analysis of MRC-ICU as a predictor of total interventions.

A total of 6 models were developed: a base model of MRC-ICU and APACHE II, full and stepwise regression, and three supervised machine learning models (see **Table 4**). All models performed similarly, while the Support Vector Machine and XGBoost models performed most consistently across all three metrics. The Random Forest model had the lowest RMSE (9.26) while Support Vector Machine had the lowest MAE (4.71). Notably, the best performing models all outperformed a parsimonious model combining severity of illness with medication regimen complexity, although these metrics do seem to give some indication as to projected CMM needs. Figure 1 depicts bee swarm plots with SHApley Additive exPlanations (SHAP) to depict the relative feature importance for the models. Regression models (see **Supplemental Tables 6-7**) and machine learning models (see **Table 3**) were also developed for intervention intensity.

**Figure 1.**
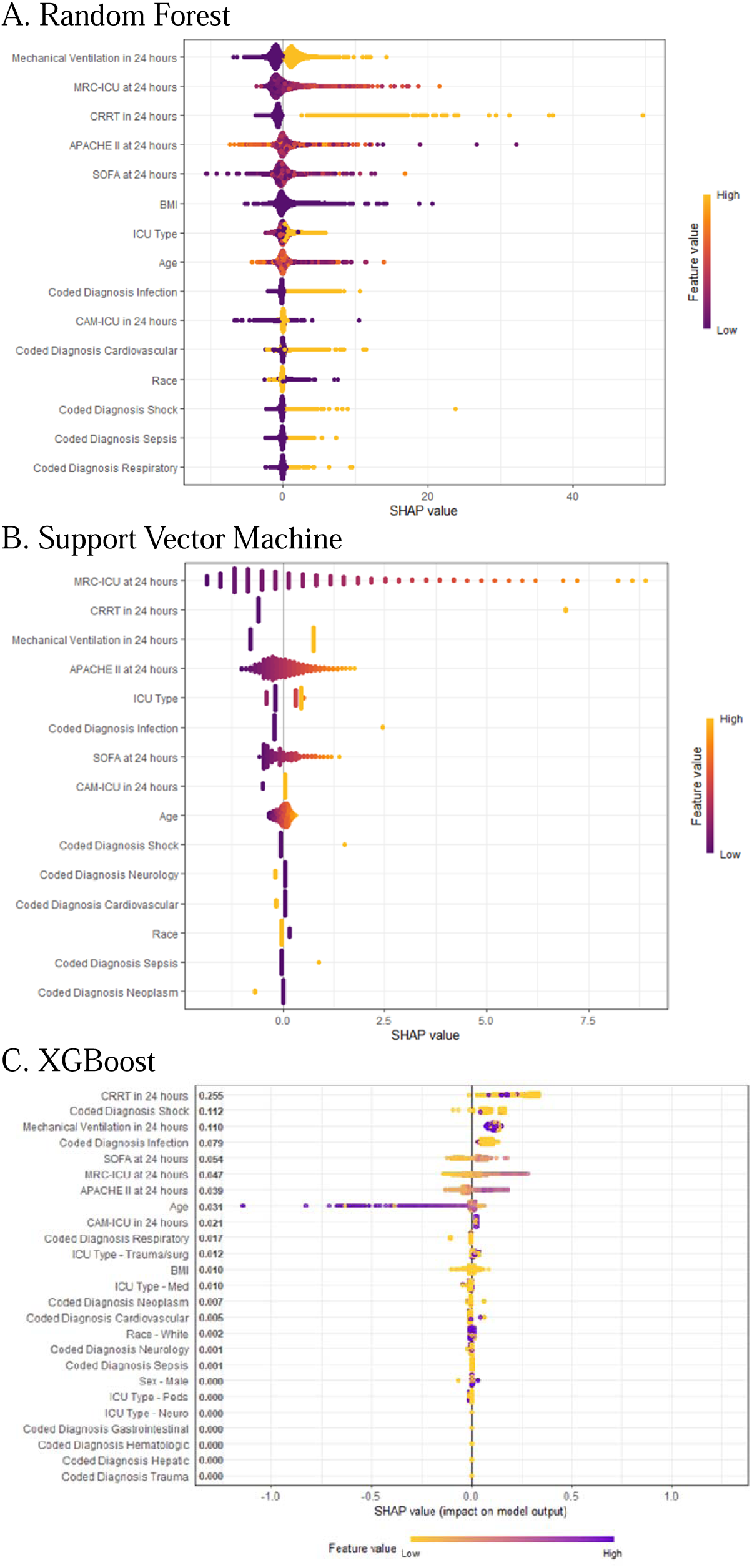
Bee Swarm Plot with SHapley Additive exPlanations (SHAP) as values for machine learning models

**Table 4.**
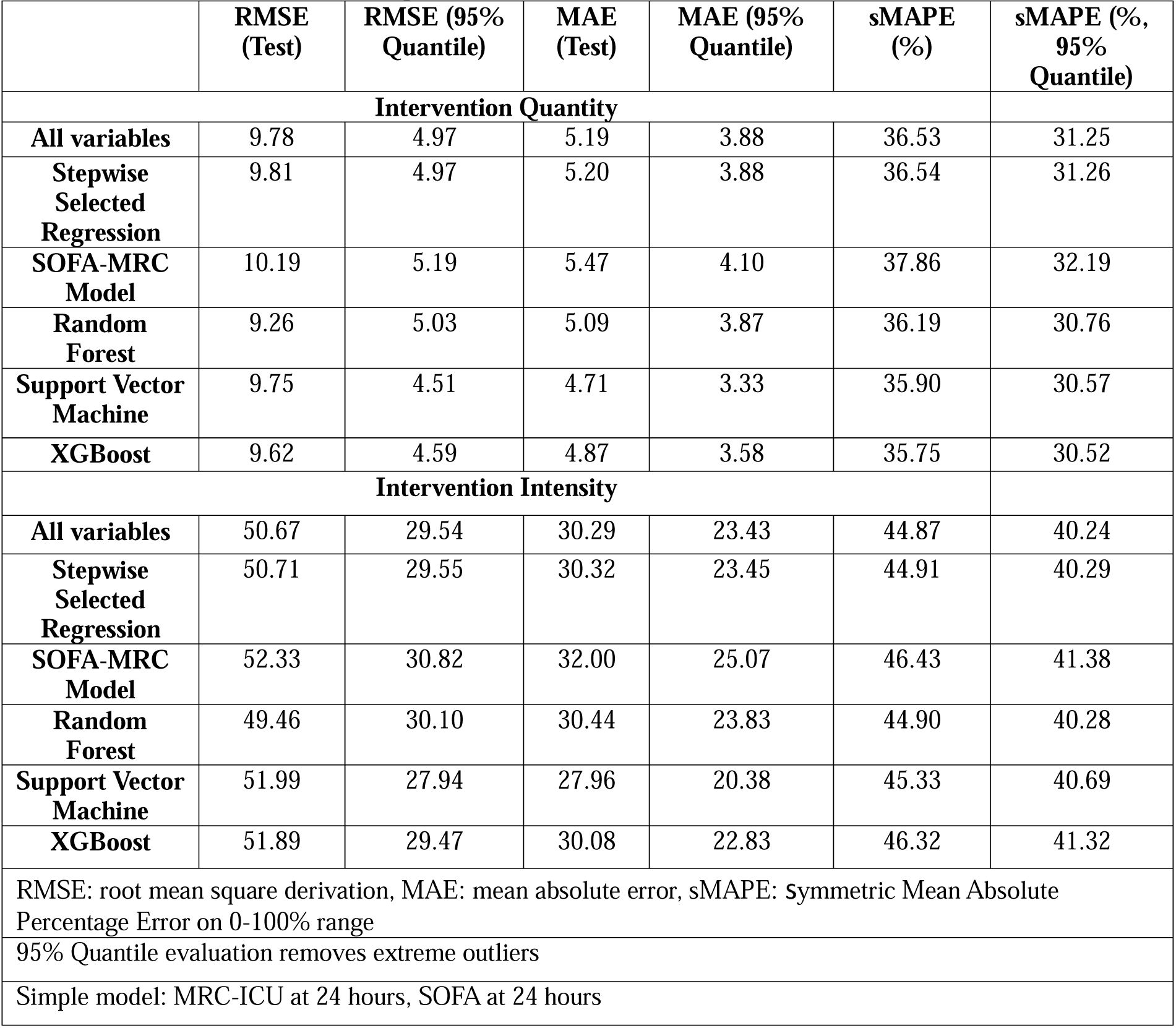
Prediction models for pharmacist interventions.

An exploratory analysis was conducted to evaluate the role of outliers in intervention quantity, given that reporting of interventions is dependent on a healthcare professional in the context of daily practice. With a 95% quantile that excluded extreme outliers, all sMAPE values were reduced, with Support Vector Machine and XGBoost performing the best at predicting interventions in the normal range and most errors due to more extreme outliers (see **Table 4**). The full regression, selected regression, and Random Forest performed better with the extreme values in their predictions (see **Appendix C**).

## Discussion

Using a specifically curated large dataset of CMM activities in critically ill patients, a series of machine learning based models were developed and found to successfully predict CMM activities, as quantified by pharmacist intervention quantity and intensity. The use of patient-specific data including traditional severity of illness scores plus CMM-oriented scores in the form of the MRC-ICU and granular pharmacist CMM activity in a large cohort marks this paper to be the first evaluation of its kind and may be useful for institutions looking to predict pharmacist workload. Performance between a stepwise regression and machine learning based models did not differ substantially; however, Random Forest and Support Vector Machine had the best performance when measured by RMSE and MAE, respectively.

The model results were reasonable, and the relationships among the covariates and outcomes were clear. Given the skewness of the intervention distributions, models showed reduced accuracy for the patients at the tail ends of intervention counts. Generally, this study suggests that severity of illness (in the form of SOFA score) and medication regimen complexity are strongly associated with critical care pharmacist interventions; however, more sophisticated models are likely necessary for improving predictive power. The similar performance between supervised machine learning methods and traditional regression has been observed in other medication related evaluations.^33, 34^ This may be due to the relatively small sample size given the heterogeneity of critically ill patients and medication use in the ICU, but also serves as an important reminder that AI based evaluations should be benchmarked against existing clinical standards and more interpretable models.

Workload optimization is important for patient and healthcare worker outcomes.^25, 35^ While cognitive overload is well known to reduce the quality of work across a variety of domains, higher patient care burdens are associated with worse patient outcomes such that standards exists for ICU physicians and nurses.^11, 25, 35–37^ The potential for additional personnel on an ‘as needed’ basis or back-up teams to be accessed for high patient care burden days in an objective and reproducible manner may be an important quality improvement strategy. CMM is a cognitive service performed with the goal to optimize medication therapy; however, a challenge to tracking such services is the lack of physical intervention.^2, 38, 39^ Reviewing a medication therapy in the context of the patient’s disease course and clinical status and comparing against current guidelines and emerging evidence is decidedly ephemeral compared to the more tangible services of central lines placed or procedures completed.^40^ Intervention tracking as a metric of CMM has been critiqued because it does not capture the cognitive labor of the process.^40^ Intervention intensity is intended to striate between lower and higher levels of such cognitive labor, but again, fails to capture times when ultimately no intervention was made, even after some amount of cognitive effort, and importantly times when a CCP made a recommendation that was not ultimately accepted by the ICU team.^29^ As such, prediction models that track intervention quantity or intensity cannot capture this element. However, they do have the potential to serve as markers of workload or potentially unsafe care processes that require improvement. For example, repeated interventions related to the use of neuromuscular blockers may open a discussion among ICU clinicians/leaders regarding standardized order set development.

Limitations of our evaluation include that it is a retrospective and single center study, precluding causal assessment and external validation. However, high quality, granular datasets with both CMM data, medication data, and patient data are not widely available, and this evaluation provides a novel foundation for future curation of these valuable datasets. While external validation is a gold standard for model building and testing, there is a high rate of practice variation in how institutions record pharmacist CMM activity such that readily comparable datasets are not available. However, steps were taken to increase the generalizability our results, notably through the re-coding of intervention categories into previously published categories supported by a systematic review and a previously used intensity scoring system. This methodology supports future investigations into external validation.

## Conclusion

Pharmacist interventions have a direct relationship with patient severity of illness and medication regimen complexity and may be predicted using machine learning approaches. Workload prediction has the potential to improve patient outcomes following the appropriate implementation and evaluations of such models.

## Data Availability

All data produced in the present study are available upon reasonable request to the authors

## Acknowledgements

Amoreena Most, PharmD, BCCCP; OHSU Oregon Clinical and Translational Research Institute

## Supplemental Digital Content

**Supplemental Table 1.**
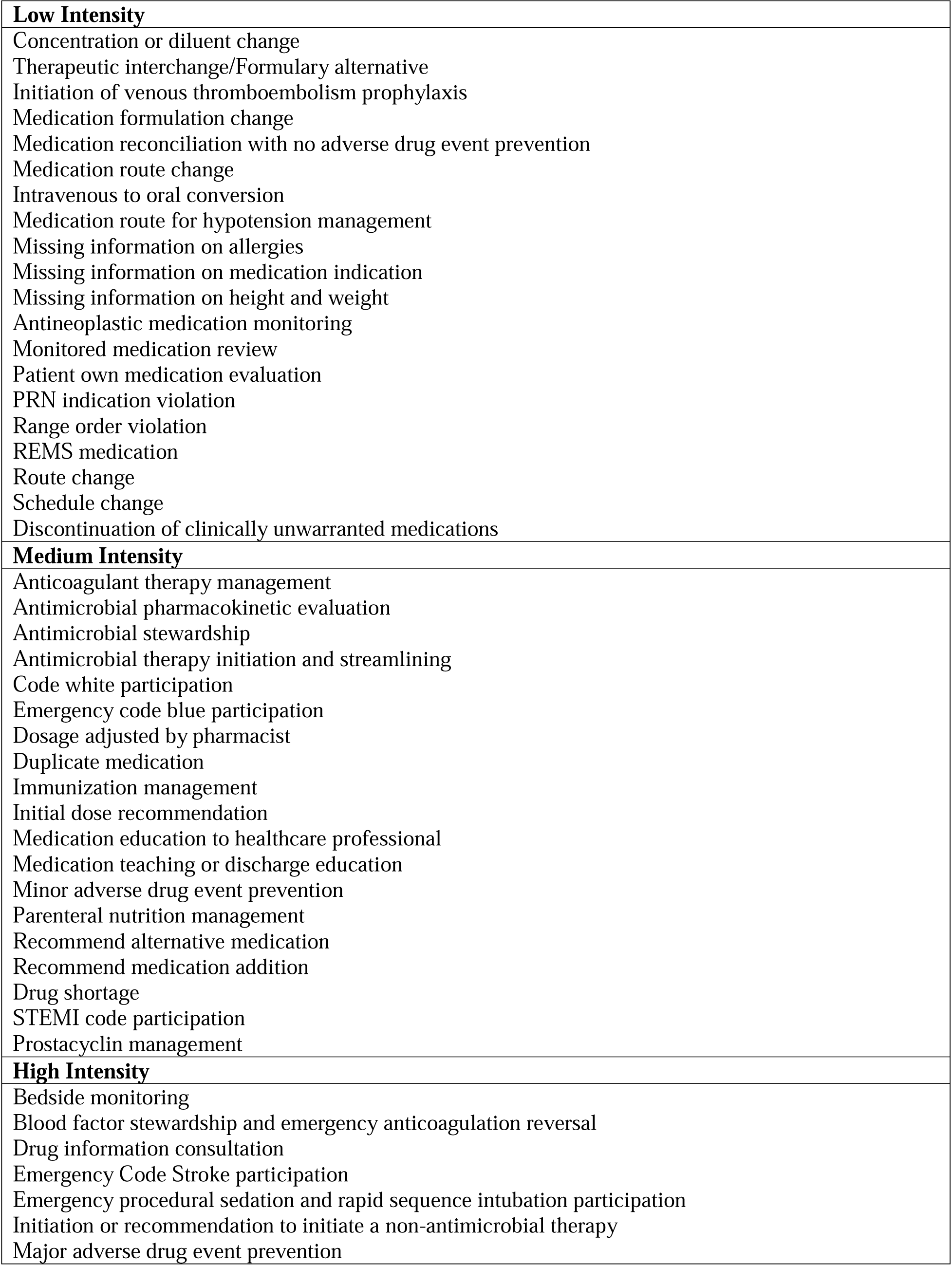
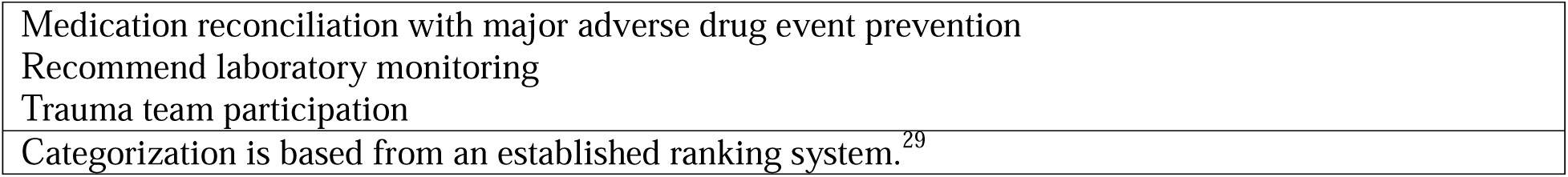
Low, medium, and high intensity intervention categories.

|  |
| --- |
| **Low Intensity** |
| Concentration or diluent change |
| Therapeutic interchange/Formulary alternative |
| Initiation of venous thromboembolism prophylaxis |
| Medication formulation change |
| Medication reconciliation with no adverse drug event prevention |
| Medication route change |
| Intravenous to oral conversion |
| Medication route for hypotension management |
| Missing information on allergies |
| Missing information on medication indication |
| Missing information on height and weight |
| Antineoplastic medication monitoring |
| Monitored medication review |
| Patient own medication evaluation |
| PRN indication violation |
| Range order violation |
| REMS medication |
| Route change |
| Schedule change |
| Discontinuation of clinically unwarranted medications |
| **Medium Intensity** |
| Anticoagulant therapy management |
| Antimicrobial pharmacokinetic evaluation |
| Antimicrobial stewardship |
| Antimicrobial therapy initiation and streamlining |
| Code white participation |
| Emergency code blue participation |
| Dosage adjusted by pharmacist |
| Duplicate medication |
| Immunization management |
| Initial dose recommendation |
| Medication education to healthcare professional |
| Medication teaching or discharge education |
| Minor adverse drug event prevention |
| Parenteral nutrition management |
| Recommend alternative medication |
| Recommend medication addition |
| Drug shortage |
| STEMI code participation |
| Prostacyclin management |
| **High Intensity** |
| Bedside monitoring |
| Blood factor stewardship and emergency anticoagulation reversal |
| Drug information consultation |
| Emergency Code Stroke participation |
| Emergency procedural sedation and rapid sequence intubation participation |
| Initiation or recommendation to initiate a non-antimicrobial therapy |
| Major adverse drug event prevention |
| Medication reconciliation with major adverse drug event prevention |
| Recommend laboratory monitoring |
| Trauma team participation |
| Categorization is based from an established ranking system.<sup>29</sup> |

**Supplemental Table 2a.** Collinearity assessment for key variables.

|  | VIF |
| --- | --- |
| Day 1 MRC-ICU | 1.626 |
| Day 1 SOFA | 1.581 |
| Day 1 APACHE II | 1.686 |
| Mechanical ventilation | 1.499 |
| CAM-ICU positive | 1.153 |
| Continuous renal replacement therapy | 1.153 |

**Supplemental Table 2b.** Correlation Matrix.

|  | Day 1 MRC-ICU | Day 1 SOFA | Day 1 APACHE II |
| --- | --- | --- | --- |
| Day 1 MRC-ICU | 1.000 | 0.451 | 0.549 |
| Day 1 SOFA | 0.451 | 1.000 | 0.466 |
| Day 1 APACHE II | 0.549 | 0.466 | 1.000 |

Co-linearity via variance inflation factor (VIF) or correlation matrix was assessed for suspected variables of interest (e.g., CRRT, mechanical ventilation, MRC-ICU, SOFA, and APACHE II score). VIF: Values above 5 indicate significant collinearity. 1 < VIF ≤ 5: Indicates moderate multicollinearity. Generally considered acceptable. VIF > 5: Indicates high multicollinearity. This suggests that the predictor variable is highly correlated with other predictors. The VIF for CRRT, Mechanical Ventilation, MRC-ICU, SOFA, APACHE all <2, which means each variable are not highly correlated with other predictors. Correlation Matrix (for continuous variables only) look for pairs of predictors with high correlation coefficients (e.g., above 0.8 or below −0.8). For the correlation matrix of the three continuous scores, their coefficients are all < 0.6, indicating minimal co-linearity.

**Table 3.** Characteristics of medication interventions.

| **Intervention Type** | **N = 98,988** |
| --- | --- |
| **Low intensity** |  |
| Concentration or diluent change | 1073 (1.0) |
| Therapeutic interchange/Formulary alternative | 399 (0.4) |
| Initiation of venous thromboembolism prophylaxis | 1092 (1.1) |
| Medication formulation change | 2979 (3.0) |
| Medication reconciliation with no adverse drug event prevention | 1877 (1.9) |
| Medication route change | 22 (0.1) |
| Medication formulation change |  |
| Medication reconciliation with no adverse drug event prevention | 4284 (4.3) |
| Missing information on allergies | 650 (0.6) |
| Missing information on medication indication | 1065 (1.0) |
| Missing information on height and weight | 520 (0.5) |
| Antineoplastic medication monitoring | 179 (0.1) |
| Monitored medication review | 2643 (2.6) |
| Patient own medication evaluation | 777 (0.7) |
| PRN indication violation | 280 (0.2) |
| Range order violation | 1035 (1.0) |
| REMS medication | 32 (0.0) |
| Route change | 9570 (0.6) |
| Schedule change | 7344 (7.4) |
| Discontinuation of clinically unwarranted medications | 13270 (13.4) |
| **Moderate intensity** |  |
| Anticoagulant therapy management | 4691 (4.7) |
| Antimicrobial pharmacokinetic evaluation | 4778 (4.8) |
| Antimicrobial stewardship | 44 (0.0) |
| Antimicrobial therapy initiation and streamlining | 1319 (1.3) |
| Code white participation | 63 (0.1) |
| Emergency code blue participation | 111 (0.1) |
| Dosage adjusted by pharmacist | 11985 (12.1) |
| Duplicate medication | 5045 (5.1) |
| Immunization management | 274 (0.2) |
| Initial dose recommendation | 2377 (2.4) |
| Medication education to healthcare professional | 2983 (3.0) |
| Medication teaching or discharge education | 1567 (1.5) |
| Minor adverse drug event prevention | 783 (0.7) |
| Parenteral nutrition management | 677 (0.6) |
| Recommend alternative medication | 2156 (2.1) |
| Recommend medication addition | 592 (0.6) |
| Drug shortage | 103 (0.1) |
| STEMI code participation | 15 (0.1) |
| Prostacyclin management | 18 (0.1) |
| **High intensity** |  |
| Bedside monitoring | 37 (0.1) |
| Blood factor stewardship and emergency anticoagulation reversal | 61 (0.1) |
| Drug information consultation | 594 (0.6) |
| Emergency Code Stroke participation | 43 (0.1) |
| Emergency procedural sedation and rapid sequence intubation participation | 28 (0.1) |
| Initiation or recommendation to initiate a non-antimicrobial therapy | 5960 (6.0) |
| Major adverse drug event prevention | 93 (0.9) |
| Medication reconciliation with major adverse drug event prevention | 1228 (1.2) |
| Recommend laboratory monitoring | 2058 (2.0) |
| Trauma team participation | 224 (0.2) |
| Data are presented as n(%) unless otherwise marked. |  |

**Supplemental Table 4.**
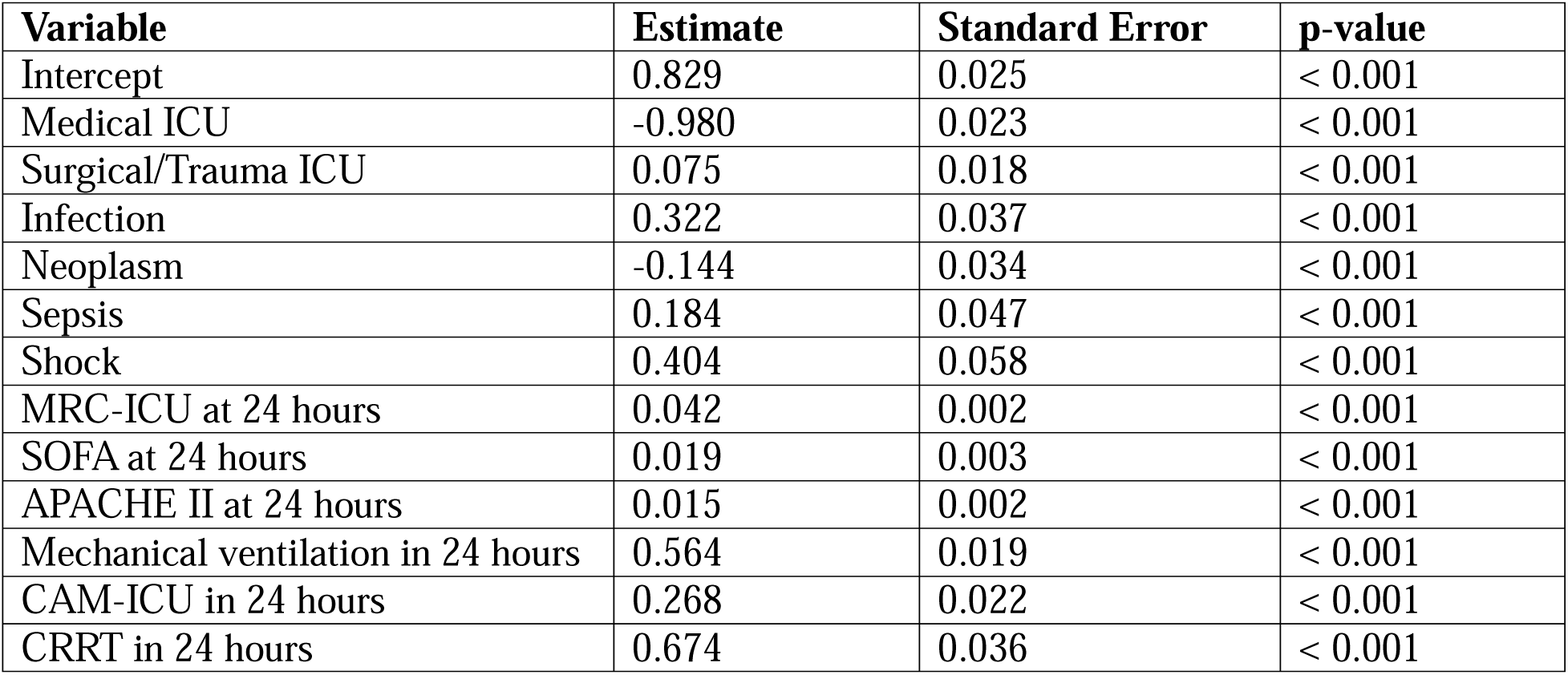
Final regression model for prediction model of total number of interventions.

| **Variable** | **Estimate** | **Standard Error** | **p-value** |
| --- | --- | --- | --- |
| Intercept | 0.829 | 0.025 | < 0.001 |
| Medical ICU | -0.980 | 0.023 | < 0.001 |
| Surgical/Trauma ICU | 0.075 | 0.018 | < 0.001 |
| Infection | 0.322 | 0.037 | < 0.001 |
| Neoplasm | -0.144 | 0.034 | < 0.001 |
| Sepsis | 0.184 | 0.047 | < 0.001 |
| Shock | 0.404 | 0.058 | < 0.001 |
| MRC-ICU at 24 hours | 0.042 | 0.002 | < 0.001 |
| SOFA at 24 hours | 0.019 | 0.003 | < 0.001 |
| APACHE II at 24 hours | 0.015 | 0.002 | < 0.001 |
| Mechanical ventilation in 24 hours | 0.564 | 0.019 | < 0.001 |
| CAM-ICU in 24 hours | 0.268 | 0.022 | < 0.001 |
| CRRT in 24 hours | 0.674 | 0.036 | < 0.001 |

**Supplemental Table 5.** Simple prediction model for total number of interventions.

| Variable | Estimate<br>(Standard Error) | Rate Ratio (95%<br>Confidence Interval) | p-value |
| --- | --- | --- | --- |
| Intercept | 1.240 (1.211) | - | < 0.001 |
| MRC-ICU at 24 hours | 0.079 (0.075) | 1.083 (0.075) | < 0.001 |
| SOFA at 24 hours | 0.072 (0.066) | 1.075 (0.066) | < 0.001 |

**Supplemental Table 6.**
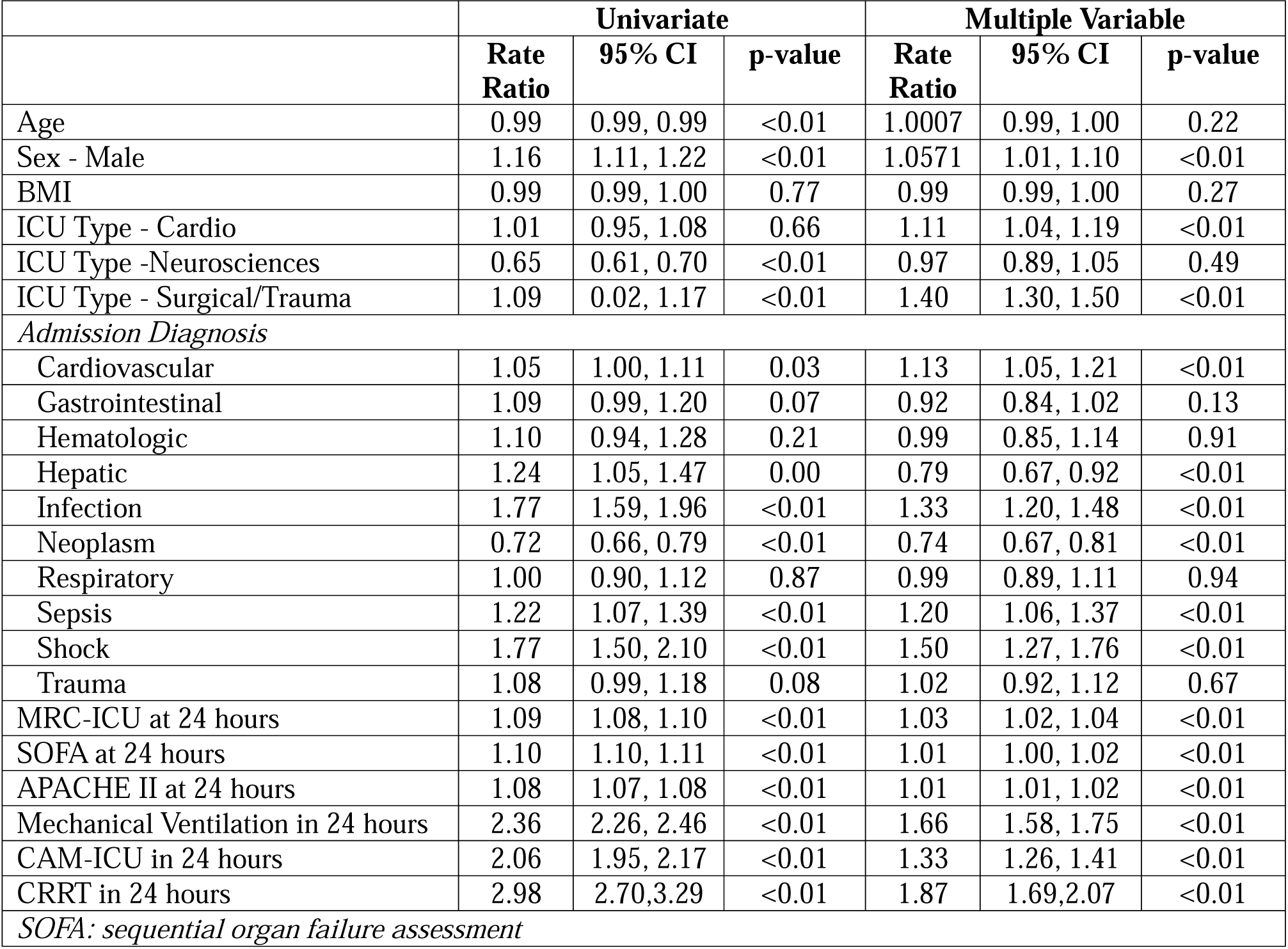
Univariate and multivariable analysis of MRC-ICU as a predictor of intervention intensity.

**Supplemental Table 7.** Final regression model for prediction model for interventions intensity.

| Variable | Estimate | Standard Error | p-value |
| --- | --- | --- | --- |
| Intercept | 2.495 | 0.040 | <0.001 |
| ICU type - Medical | -0.125 | 0.035 | <0.001 |
| ICU type - Neuroscience | -0.167 | 0.035 | <0.001 |
| ICU type - Surgical/Trauma | 0.224 | 0.032 | <0.001 |
| Cardiovascular | 0.152 | 0.033 | <0.001 |
| Infection | 0.321 | 0.051 | <0.001 |
| Neoplasm | -0.268 | 0.045 | <0.001 |
| Sepsis | 0.212 | 0.064 | <0.001 |
| Shock | 0.431 | 0.081 | <0.001 |
| MRC-ICU at 24 hours | 0.035 | 0.003 | <0.001 |
| SOFA at 24 hours | 0.014 | 0.004 | <0.001 |
| APACHE II at 24 hours | 0.016 | 0.003 | <0.001 |
| Mechanical Ventilation in 24 hours | 0.510 | 0.025 | <0.001 |
| CAM-ICU in 24 hours | 0.290 | 0.029 | <0.001 |
| CRRT in 24 hours | 0.610 | 0.050 | <0.001 |

**Supplemental Table 8.** Missing Data.

| Number of Missing Values |  |
| --- | --- |
| Age | 13 |
| Female | 0 |
| Race | 349 |
| BMI | 741 |
| ICU Type | 0 |
| Cardiovascular | 0 |
| Gastrointestinal | 0 |
| Hematologic | 0 |
| Hepatic | 0 |
| Infection | 0 |
| Neoplasm | 0 |
| Neurology | 0 |
| Respiratory | 0 |
| Sepsis | 0 |
| Shock | 0 |
| Trauma | 0 |
| MRC-ICU at 24 hours | 683 |
| SOFA at 24 hours | 208 |
| APACHE II at 24 hours | 13 |
| Mechanical Ventilation in 24 hours | 0 |
| CAM-ICU in 24 hours | 0 |
| CRRT in 24 hours | 0 |
| Total in-hospital interventions | 0 |
| Overall composite intervention score | 0 |

**Supplemental Digital Content Figure 1.**
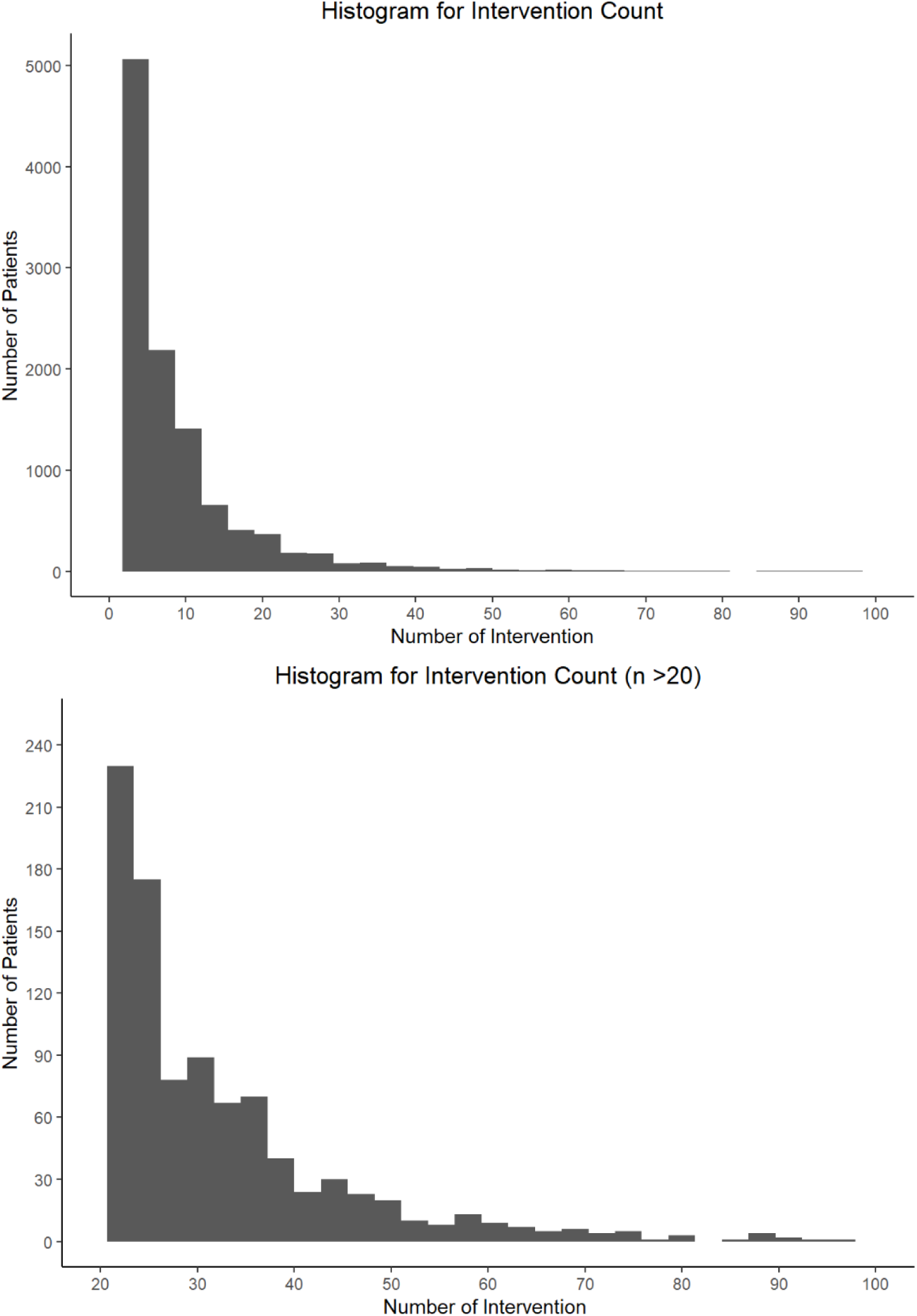
Histogram of intervention counts Top panel: Histogram of number of interventions by patient. Bottom panel: Histogram with
amplified tail for clearer visualization of patients with greater than 20 interventions.

**Supplemental Digital Content Figure 2.**
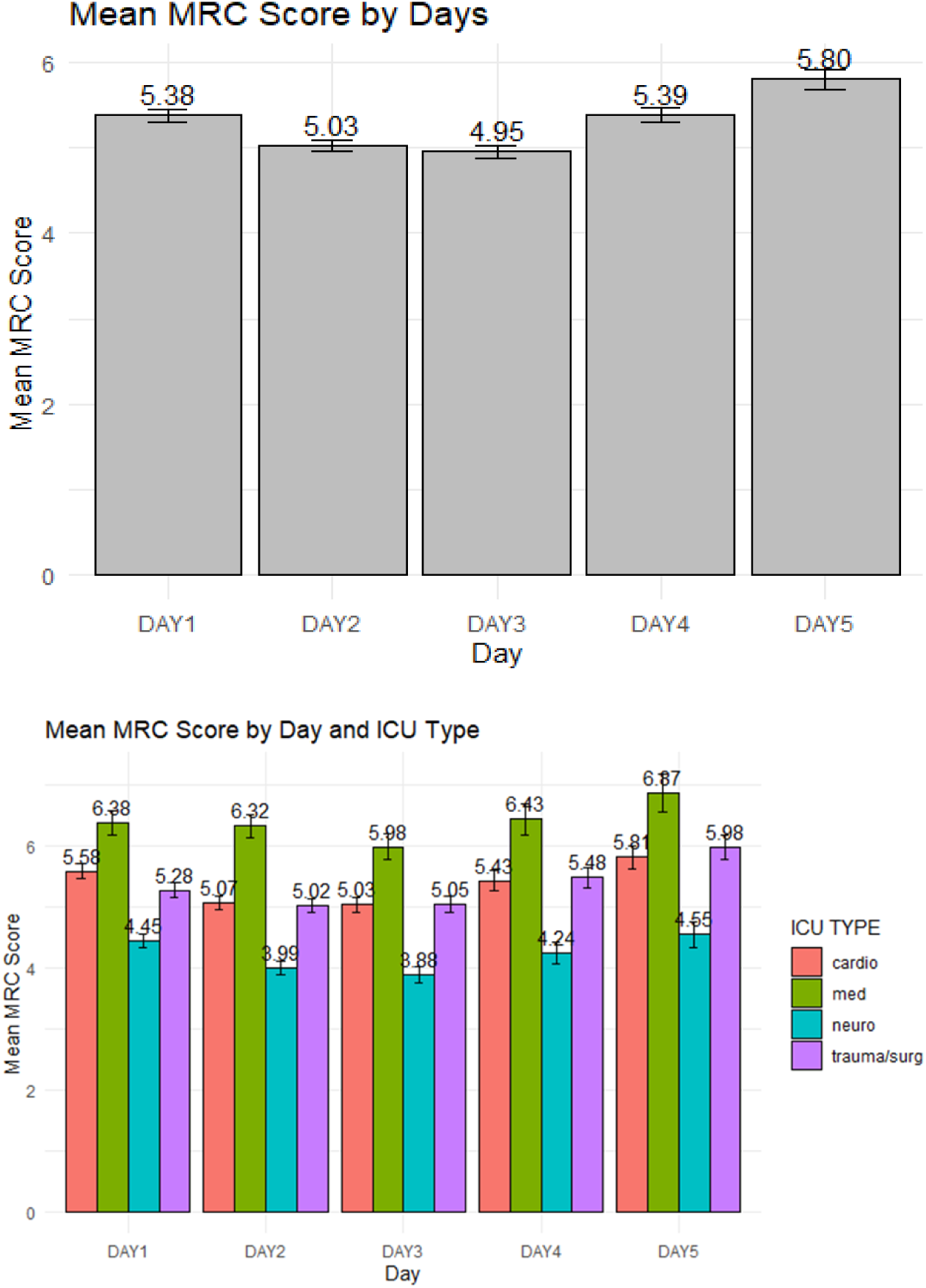
MRC-IRU Score by ICU Day

**Supplemental Digital Content Figure 3.**
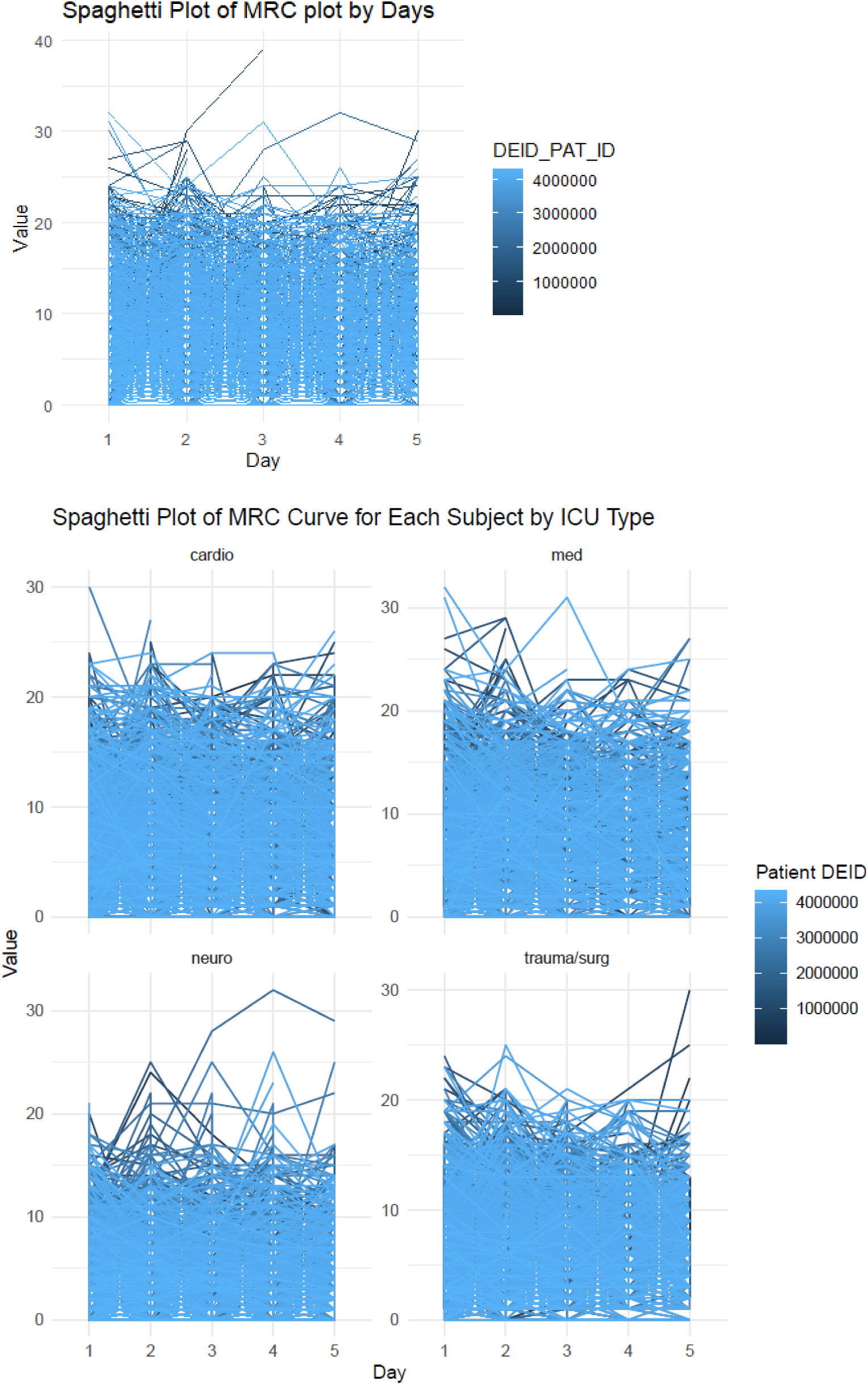
MRC-ICU changes over ICU day

**Supplemental Digital Content Figure 4.**
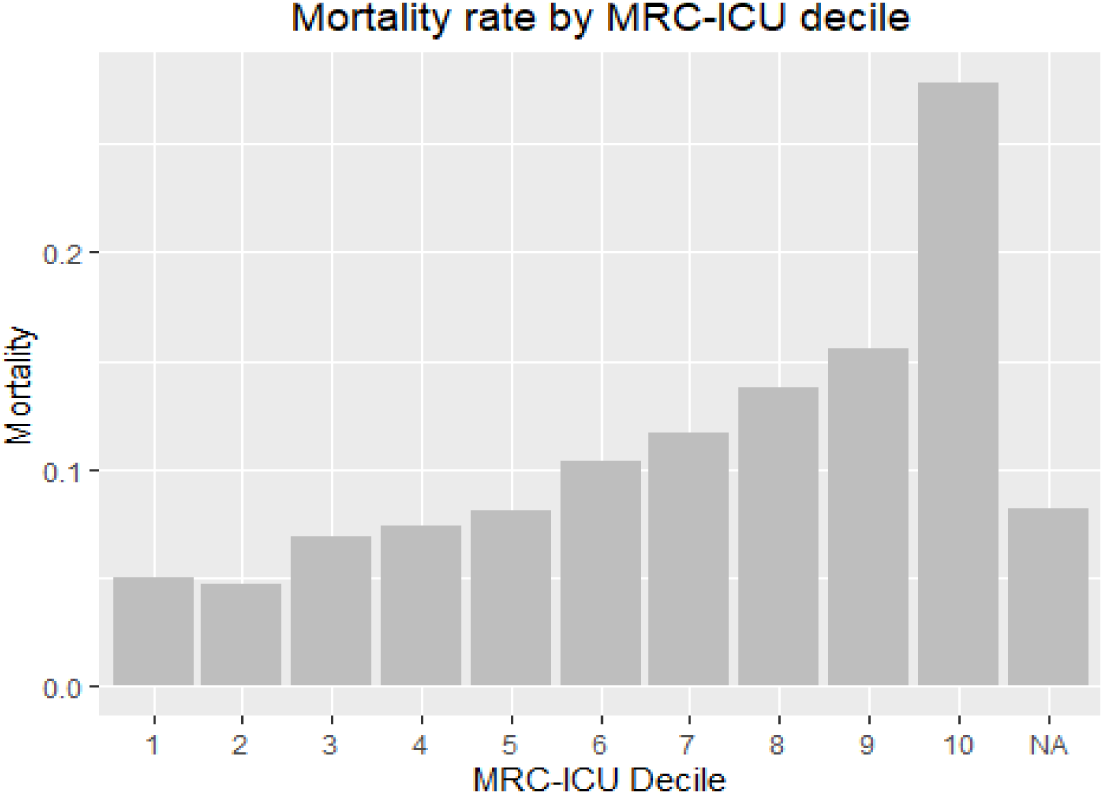
Mortality rate by MRC-IRU decile

**Supplemental Digital Content Figure 5.**
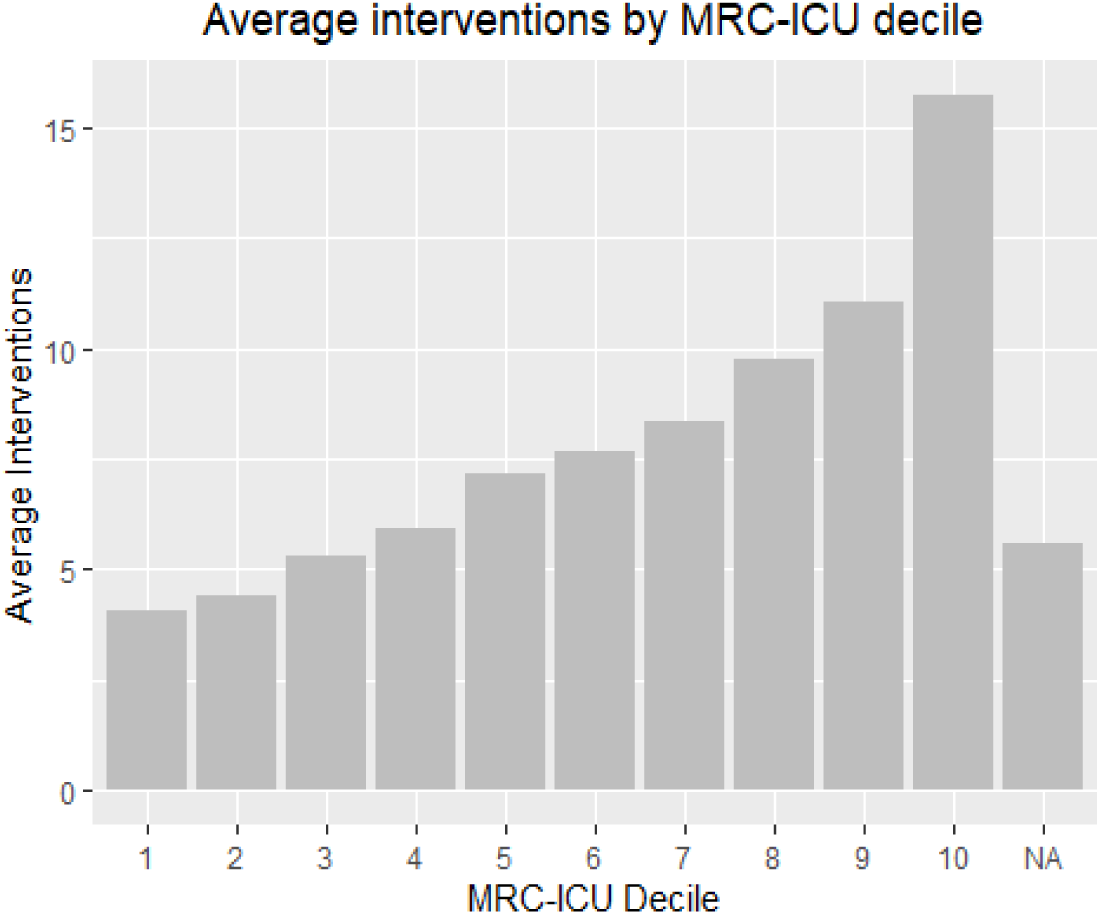
Interventions by MRC-ICU decile

**Supplemental Figure 6.**
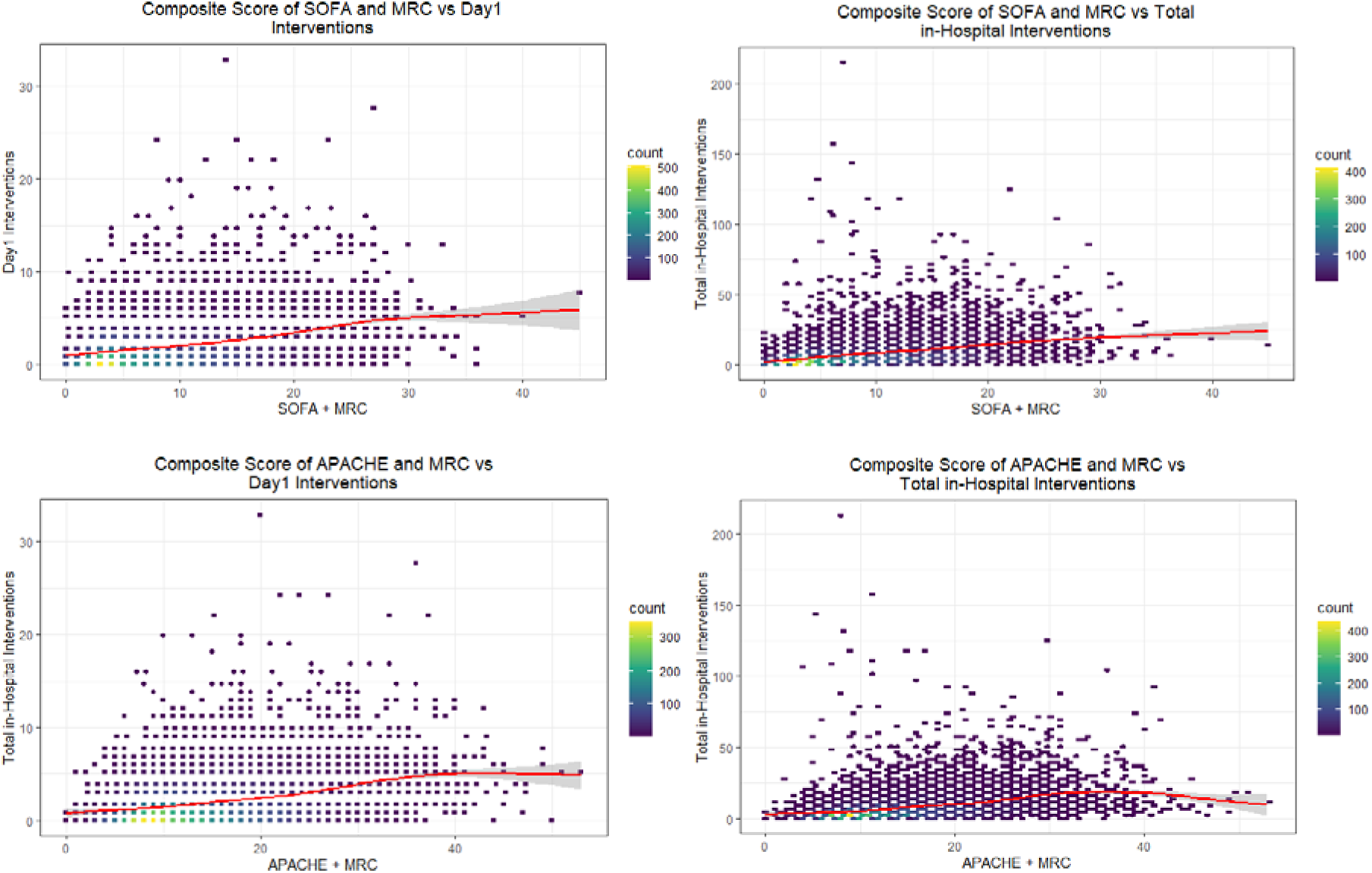
Composite severity of illness and medication data with pharmacist interventions

**Supplemental Figure 7.**
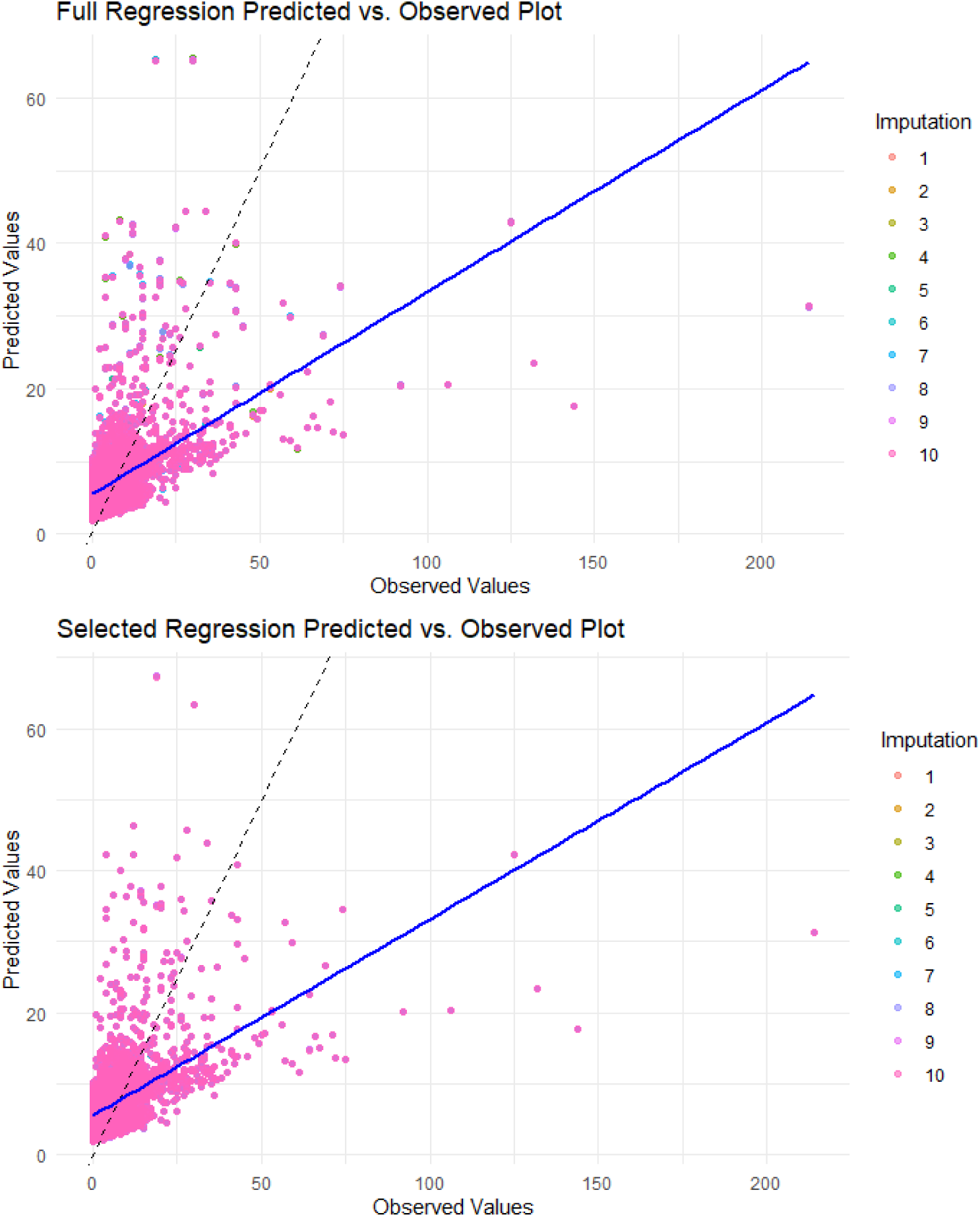
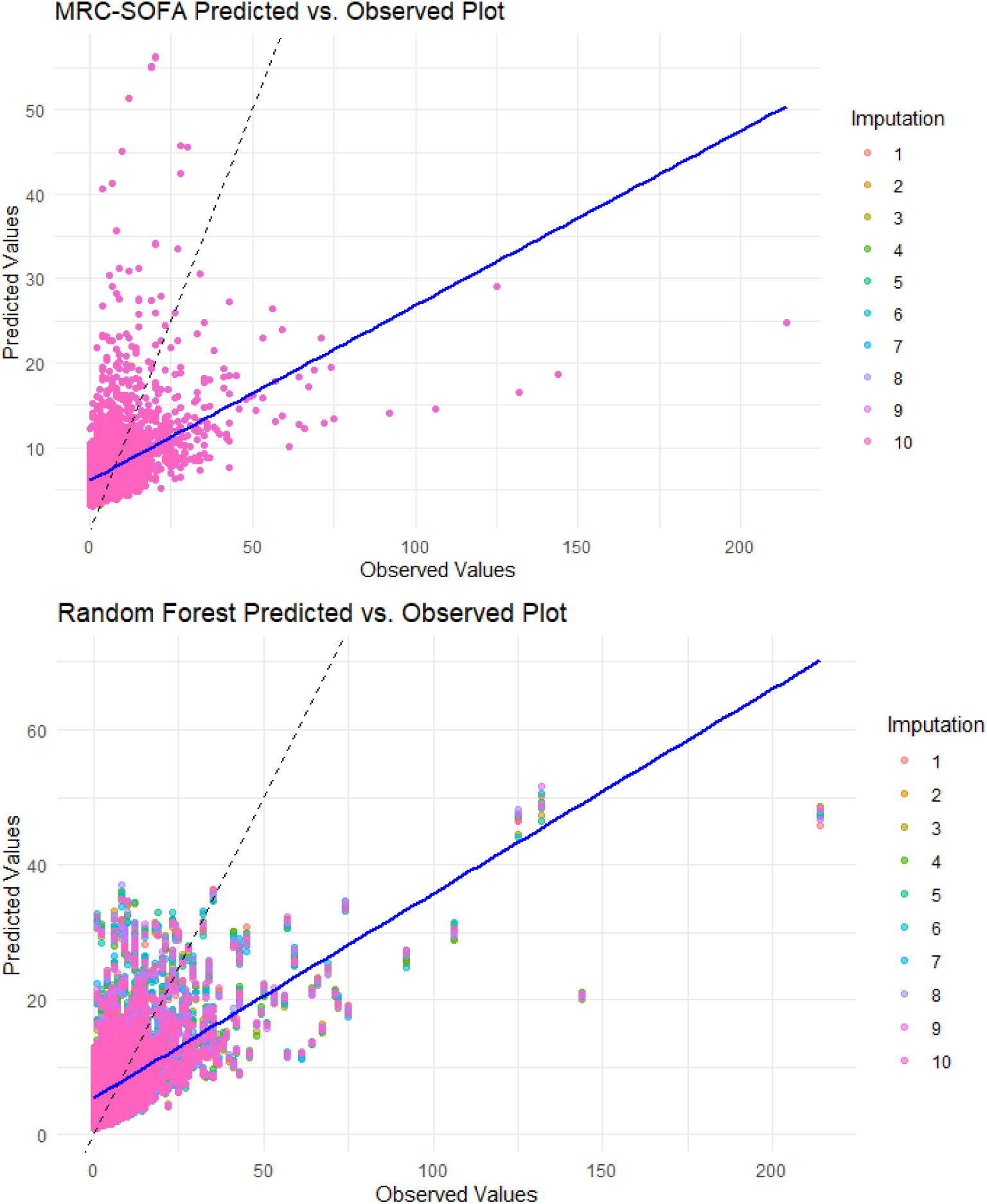
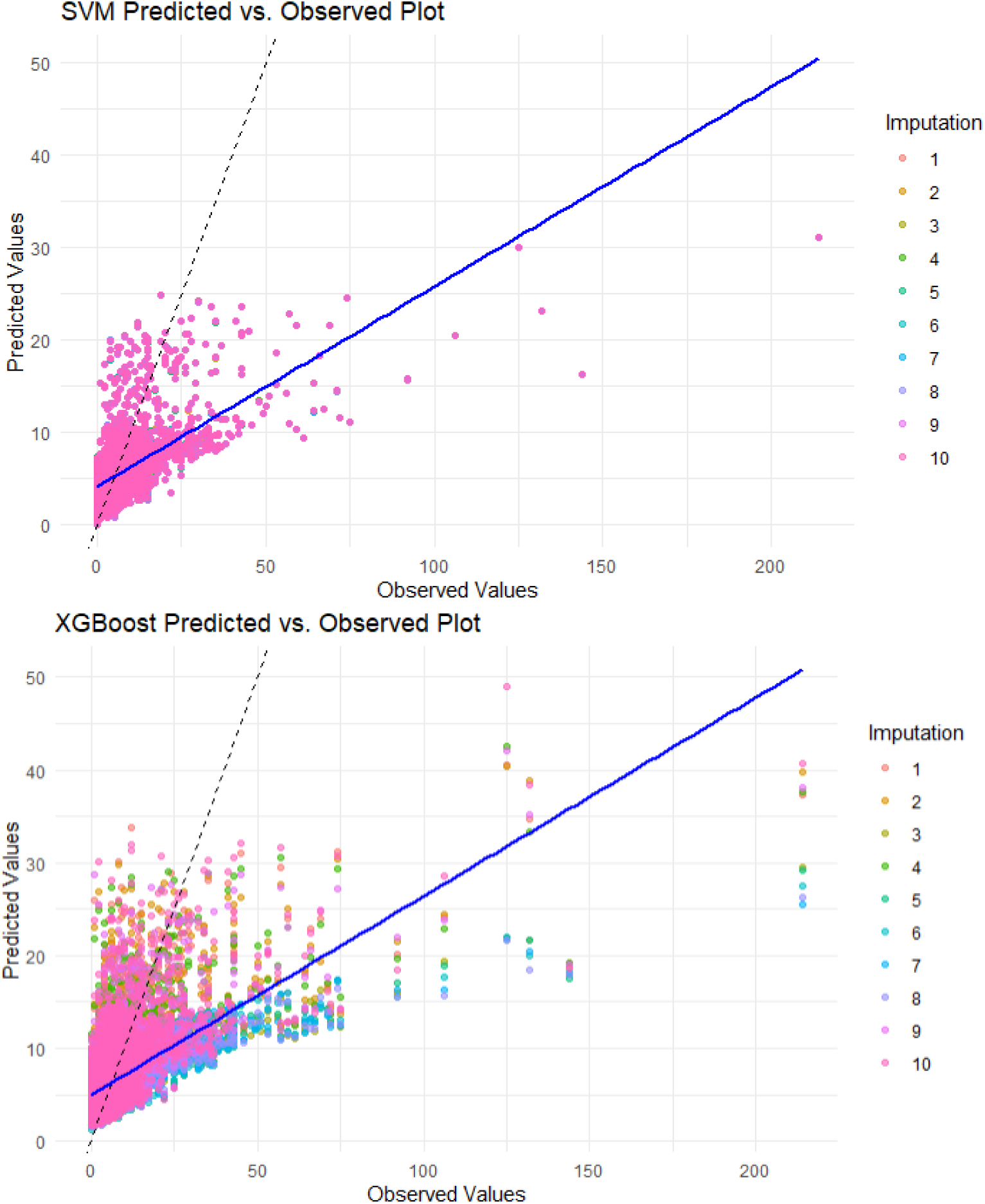
Predicted Intervention VS Observed Interventions for each Models

## Appendix A. Model Selection

Multivariable generalized linear models (GLM) were developed to evaluate the relationship of MRC-ICU on ICU day 1 in relation to total interventions. Both GLM with Poisson distribution and negative binomial distribution were explored. Poisson regression is a common approach when the response variable is a count like number of interventions. However, it assumes that the mean and variance of the count data are equal. This is often not the case in real-world data.

Overdispersion occurs when the observed variance is greater than the mean. An alternative approach to handle overdispersion is a negative binomial model. The negative binomial model introduces an additional parameter (the dispersion parameter) to account for the extra variability in the data. By accommodating overdispersion, the negative binomial model provides a better fit for the data, leading to more reliable estimates and inference. Because the variance of the total intervention (93.22) was much greater than its mean (7.40), and the Pearson statistics from full multivariate Poisson regression without missing imputation also indicated overdispersion (Pearson statistics=95076, df=11676, p-value<0.001), the Negative Binomial regression model was selected as a substitute for the Poisson regression. Similarly, because the variance of the intervention intensity score (2774.91) was much greater than its mean (37.68), and the Pearson statistics from full multivariate Poisson regression without missing imputation also indicated overdispersion (Pearson statistics=620060, df=11676, p-value<0.001), the Negative Binomial regression model was selected in substitute of the Poisson regression.

## Appendix B. Temporal Analysis

### Mixed Model on Daily Effect

The daily or temporal time effect of SOFA score and MRC-ICU score on the daily number of interventions was analyzed. One unit higher in MRC-ICU score was associated with 10.6% increase in number of interventions at baseline. There was no significant effect of SOFA score on number of interventions at baseline. Compared with the previous day, the effect of MRC score on number of interventions is 0.5% lower, but the effect of SOFA score on number of interventions is 1.5% higher.

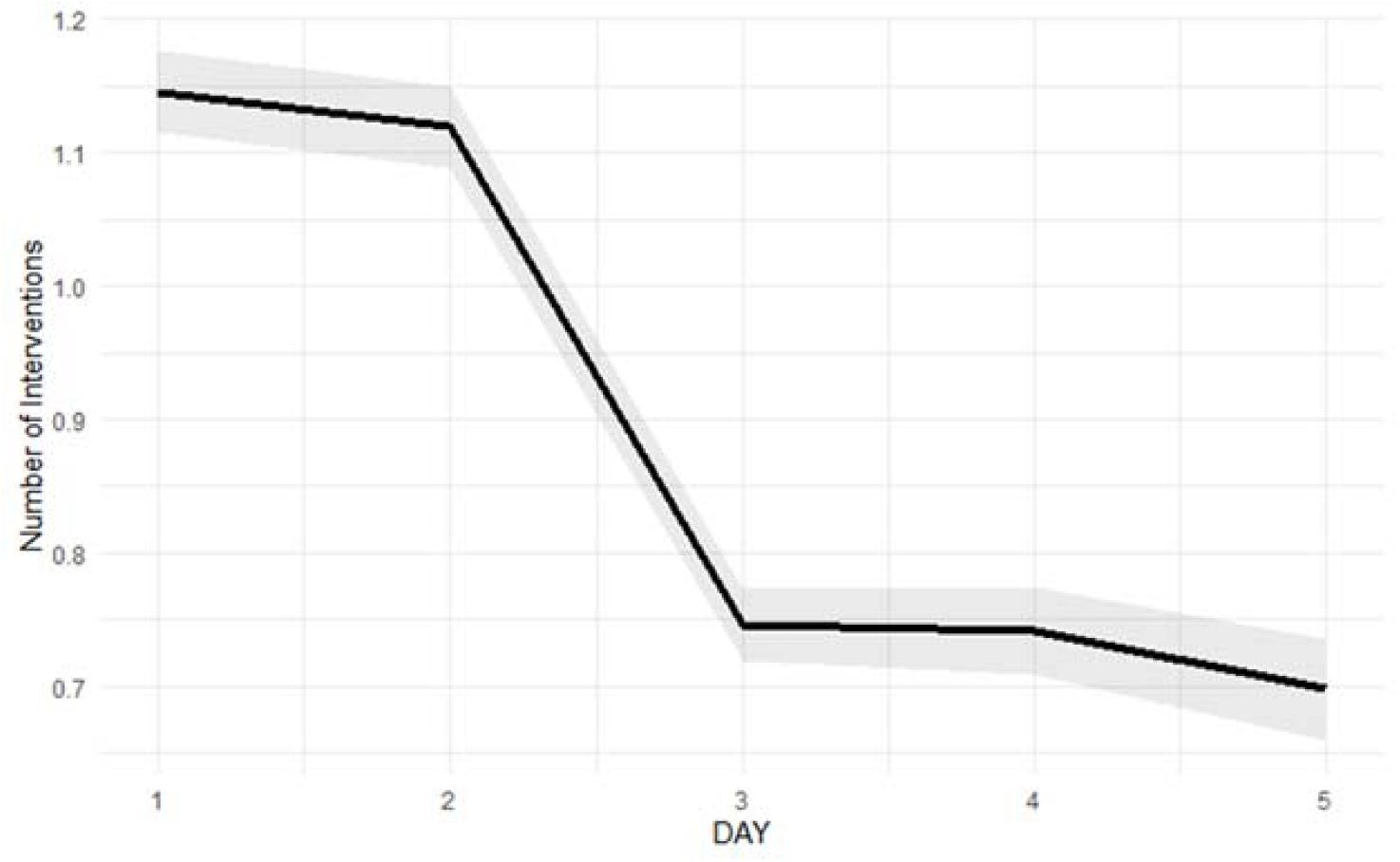

| Variable | Relative Risk | 95% Confidence Interval | p-value |
| --- | --- | --- | --- |
| Intercept | 0.537 | 0.490-0.590 | 0.000 |
| Age | 1.000 | 0.999-1.001 | 0.420 |
| Male sex | 1.002 | 0.969-1.036 | 0.915 |
| Medical ICU | 0.904 | 0.861-0.949 | 0.000 |
| Trauma/Surgical ICU | 1.336 | 1.287-1.388 | 0.000 |
| APACHE at 24 hours | 1.014 | 1.010-1.019 | 0.000 |
| Infection | 1.002 | 0.930-1.081 | 0.949 |
| Neoplasm | 0.900 | 0.836-0.968 | 0.005 |
| Neurology | 1.120 | 1.073-1.169 | 0.000 |
| Sepsis | 1.091 | 0.993-1.200 | 0.071 |
| Shock | 1.317 | 1.176-1.475 | 0.000 |
| SOFA score | 0.997 | 0.985-1.008 | 0.560 |
| MRC-ICU day 1 | 1.106 | 1.097-1.116 | 0.000 |
| Day | 0.832 | 0.815-0.849 | 0.000 |
| SOFA Score: Day | 1.015 | 1.011-1.019 | 0.000 |
| MRC-ICU: Day | 0.995 | 0.992-0.998 | 0.002 |

## Appendix C. Subgroup analysis

**Table 1b.** Subgroup analysis for number of interventions. The overall performance of each model was similar, but the mean and variance for each model was quite different (e.g., mean for full regression prediction was 7.58 while mean for SVM prediction was 5.47). A subgroup analysis for low, medium, and high intervention groups was conducted (Table 1a-b), and distribution of errors were plotted for number of interventions (Figure 1a) and intervention intensity score (Figure 1b).

| Table 1a. Subgroup analysis for number of interventions |  |  |  |  |  |
| --- | --- | --- | --- | --- | --- |
| | Predicted Mean (SD) | True Mean (SD) | RMSE for low intervention group (<5) | RMSE for medium intervention group ($\geq 5$, <10) | RMSE for high intervention group ($\geq 10$) |
| Full Regression | 7.58 (5.61) | 7.49 (10.44) | 4.91 | 4.60 | 18.26 |
| Selected Regression | 7.59 (5.63) | 7.49 (10.44) | 4.91 | 4.60 | 18.32 |
| Simple Model | 7.62 (4.78) | 7.49 (10.44) | 5.04 | 4.72 | 19.09 |
| Random Forest | 7.72 (5.25) | 7.49 (10.44) | 5.05 | 4.91 | 16.85 |
| SVM | 5.47 (3.49) | 7.49 (10.44) | 3.15 | 3.23 | 19.31 |
| XGBoost | 6.50 (3.15) | 7.49 (10.44) | 3.95 | 2.93 | 18.54 |
| Table 1b. Subgroup analysis for intervention intensity |  |  |  |  |  |
| | Predicted Mean (SD) | True Mean (SD) | RMSE for low intervention intensity score group (<30) | RMSE for medium intervention intensity score group ($\geq 30$, <60) | RMSE for high intervention intensity score group ($\geq 60$) |
| Full Regression | 38.33 (26.31) | 38.57 (54.10) | 27.90 | 24.31 | 101.20 |
| Selected Regression | 38.30 (26.08) | 38.57 (54.10) | 27.89 | 24.56 | 101.22 |
| Simple Model | 38.40 (20.38) | 38.57 (54.10) | 28.18 | 22.58 | 105.68 |
| Random Forest | 39.80 (26.45) | 38.57 (54.10) | 29.55 | 26.32 | 95.86 |
| SVM | 24.62 (16.31) | 38.57 (54.10) | 16.34 | 22.47 | 112.09 |
| XGBoost | 30.79 (14.04) | 38.57 (54.10) | 20.51 | 17.38 | 107.75 |

**Figure 1a.**
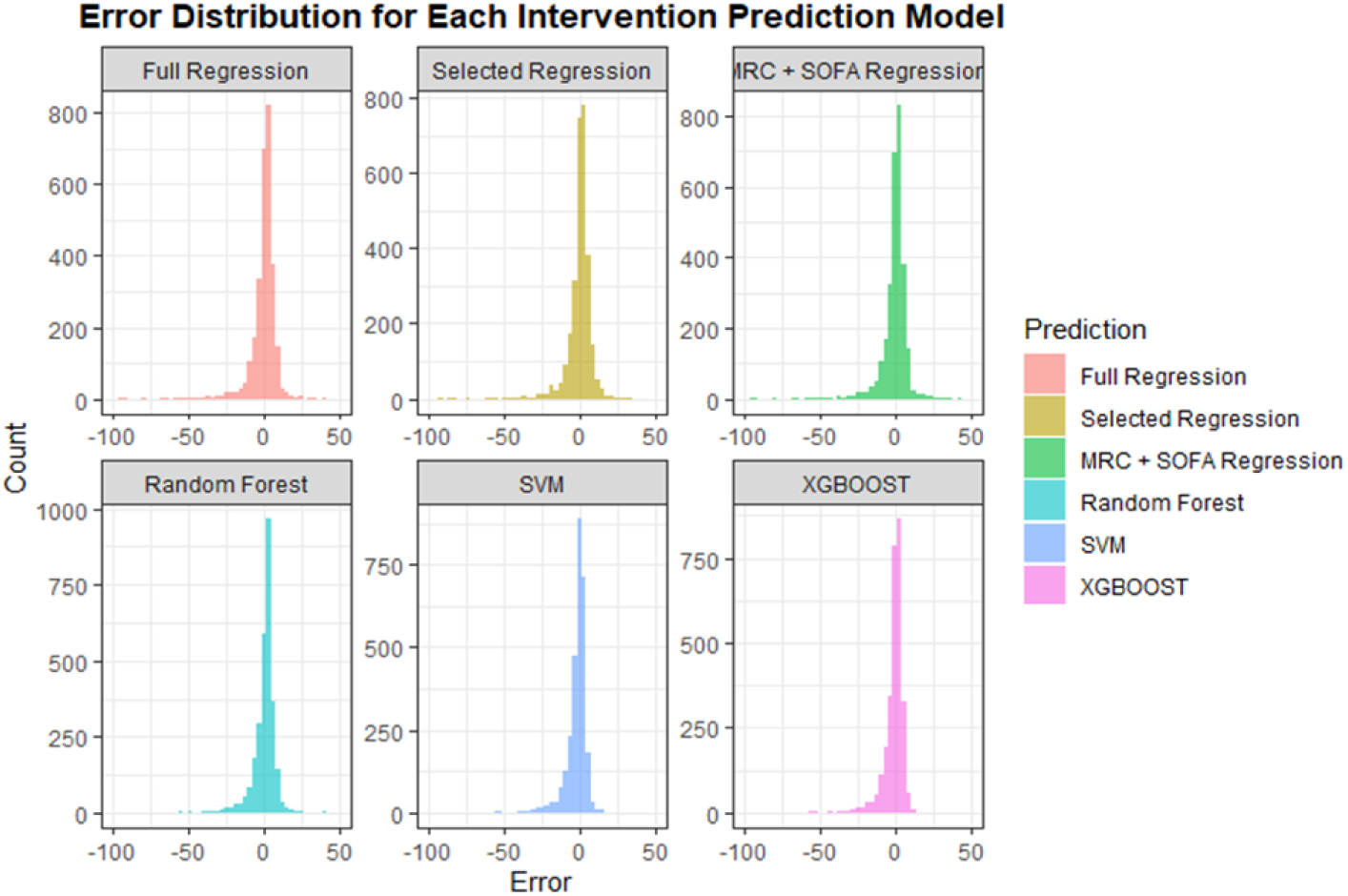
Error distribution for number of interventions

**Figure 1b.**
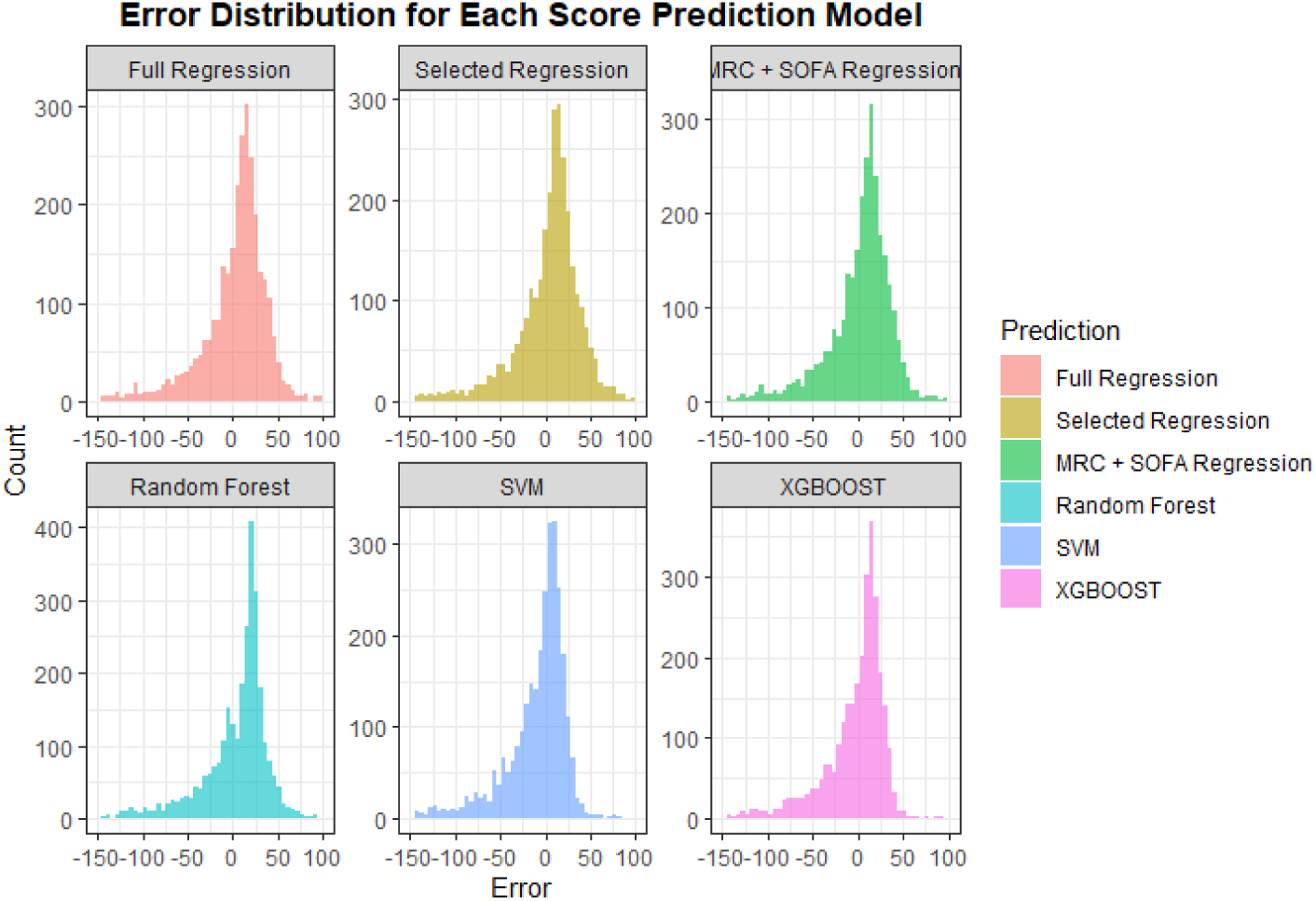
Error distribution for intervention intensity score

## References

1. A S. Critical Care Pharmacists: A Focus on Horizons. Critical Care Clinics 2023;3:503–27.

2. Newsome AS, Murray B, Smith SE, et al. Optimization of critical care pharmacy clinical services: A gap analysis approach. Am J Health Syst Pharm 2021;22:2077–85.

3. Halpern NA, Goldman DA, Tan KS, Pastores SM. Trends in Critical Care Beds and Use Among Population Groups and Medicare and Medicaid Beneficiaries in the United States: 2000-2010. Crit Care Med 2016;8:1490–9.

4. Cullen DJ, Sweitzer BJ, Bates DW, Burdick E, Edmondson A, Leape LL. Preventable adverse drug events in hospitalized patients: a comparative study of intensive care and general care units. Crit Care Med 1997;8:1289–97.

5. Practices IoSM. High Alert Medications Available from https://www.ismp.org/sites/default/files/attachments/2018-08/highAlert2018-Acute-Final.pdf.

6. Maslove DM, Lamontagne F, Marshall JC, Heyland DK. A path to precision in the ICU. Crit Care 2017;1:79.

7. Shulman R, McKenzie CA, Landa J, et al. Pharmacist’s review and outcomes: Treatment-enhancing contributions tallied, evaluated, and documented (PROTECTED-UK). J Crit Care 2015;4:808–13.

8. Lee H, Ryu K, Sohn Y, Kim J, Suh GY, Kim E. Impact on Patient Outcomes of Pharmacist Participation in Multidisciplinary Critical Care Teams: A Systematic Review and Meta-Analysis. Crit Care Med 2019;9:1243–50.

9. Leape LL, Cullen DJ, Clapp MD, et al. Pharmacist participation on physician rounds and adverse drug events in the intensive care unit. JAMA 1999;3:267–70.

10. MacLaren R, Roberts RJ, Dzierba AL, Buckley M, Lat I, Lam SW. Characterizing Critical Care Pharmacy Services Across the United States. Crit Care Explor 2021;1:e0323.

11. Newsome AS, Smith SE, Jones TW, Taylor A, Van Berkel MA, Rabinovich M. A survey of critical care pharmacists to patient ratios and practice characteristics in intensive care units. JACCP: JOURNAL OF THE AMERICAN COLLEGE OF CLINICAL PHARMACY 2020;1:68–74.

12. Smetana KS, Flannery AH, Gurnani PK, et al. PHarmacist avoidance or reductions in medical costs in CRITically ill adults rounding with one SERVICE compared to two or more services: PHARM-CRIT-SERVICE. JACCP: JOURNAL OF THE AMERICAN COLLEGE OF CLINICAL PHARMACYn/a.

13. Sikora A, Ayyala D, Rech MA, et al. Impact of Pharmacists to Improve Patient Care in the Critically Ill: A Large Multicenter Analysis Using Meaningful Metrics With the Medication Regimen Complexity-ICU (MRC-ICU). Crit Care Med 2022.

14. Sikora A, Devlin JW, Yu M, et al. Evaluation of medication regimen complexity as a predictor for mortality. Sci Rep 2023;1:10784.

15. Gwynn ME, Poisson MO, Waller JL, Newsome AS. Development and validation of a medication regimen complexity scoring tool for critically ill patients. Am J Health Syst Pharm 2019;Supplement_2:S34-S40.

16. Olney WJ, Chase AM, Hannah SA, Smith SE, Newsome AS. Medication Regimen Complexity Score as an Indicator of Fluid Balance in Critically Ill Patients. J Pharm Pract 2022;4:573–79.

17. Rafiei A, Rad MG, Sikora A, Kamaleswaran R. Improving irregular temporal modeling by integrating synthetic data to the electronic medical record using conditional GANs: a case study of fluid overload prediction in the intensive care unit. medRxiv 2023;2023.06.20.23291680.

18. Sikora A, Zhang T, Murphy DJ, et al. Machine learning vs. traditional regression analysis for fluid overload prediction in the ICU. medRxiv 2023;2023.06.16.23291493.

19. Murray B, Zhao B, Kong Y, Shen Y, Sikora A. 935: PREDICTING DURATION OF MECHANICAL VENTILATION WITH MEDICATION REGIMEN COMPLEXITY VARIABLES. Critical Care Medicine 2023;1:460.

20. Webb AJ, Rowe S, Sikora Newsome A. A descriptive report of the rapid implementation of automated MRC-ICU calculations in the EMR of an academic medical center. Am J Health Syst Pharm 2022.

21. Sikora A, Rafiei A, Rad MG, et al. Pharmacophenotype identification of intensive care unit medications using unsupervised cluster analysis of the ICURx common data model. Crit Care 2023;1:167.

22. Chase A AH, Forehand C, Keats K, Taylor A, Wu S, Blotske K, Sikora A. An evaluation of medication regimen complexity’s relationship to medication errors in critically ill patients. Hospital Pharmacy 2023.

23. Al-Mamun MA, Brothers T, Newsome AS. Development of Machine Learning Models to Validate a Medication Regimen Complexity Scoring Tool for Critically Ill Patients. Ann Pharmacother 2021;4:421–29.

24. Newsome AS, Anderson D, Gwynn ME, Waller JL. Characterization of changes in medication complexity using a modified scoring tool. Am J Health Syst Pharm 2019;Supplement_4:S92-S95.

25. Sikora A, Ayyala D, Rech MA, et al. Impact of Pharmacists to Improve Patient Care in the Critically Ill: A Large Multicenter Analysis Using Meaningful Metrics With the Medication Regimen Complexity-ICU (MRC-ICU) Score. Crit Care Med 2022;9:1318–28.

26. Sikora A, Rafiei A, Rad MG, et al. Pharmacophenotype identification of intensive care unit medications using unsupervised cluster analysis of the ICURx common data model. Crit Care 2023;1:167.

27. Sikora A JH, Yu M, Chen X, Murray B, Kamaleswaran R. Cluster analysis driven by unsupervised latent feature learning of intensive care unit medications to identify novel pharmacophenotypes of critically ill patients. Research Square 2022.

28. Smith SE, Shelley R, Newsome AS. Medication regimen complexity vs patient acuity for predicting critical care pharmacist interventions. Am J Health Syst Pharm 2021.

29. Sikora A, Ayyala D, Rech MA, et al. Impact of Pharmacists to Improve Patient Care in the Critically Ill: A Large Multicenter Analysis Using Meaningful Metrics With the Medication Regimen Complexity-ICU (MRC-ICU) Score. Crit Care Med 2022;9:1318–28.

30. Lundberg S, Su-In Lee.. A Unified Approach to Interpreting Model Predictions. Neural Information Processing Systems 2017.

31. R. shapr: Prediction Explanation with Dependence-Aware Shapley Values, Available from https://cran.r-project.org/web/packages/shapr/index.html. 2024.

32. R. SHAPforxgboost: SHAP Plots for ‘XGBoost’, Available from https://cran.r-project.org/web/packages/SHAPforxgboost/index.html. 2024..

33. Sikora A, Zhang T, Murphy DJ, et al. Machine learning vs. traditional regression analysis for fluid overload prediction in the ICU. Sci Rep 2023;1:19654.

34. Brian Murray TZ, Xianyan Chen, Susan E. Smith, John W. Devlin, David J. Murphy, Andrea Sikora, Rishikesan Kamaleswaran the MRC-ICU Investigator Team. Augmenting mortality prediction with medication data and machine learning models. arixV https://www.medrxivorg/content/101101/2024041624305420v1. 2024

35. Lat I, Paciullo C, Daley MJ, et al. Position Paper on Critical Care Pharmacy Services: 2020 Update. Crit Care Med 2020;9:e813–e34.

36. Smith SE, Slaughter AA, Butler SA, Buckley MS, MacLaren R, Newsome AS. Examination of critical care pharmacist work activities and burnout. JACCP: JOURNAL OF THE AMERICAN COLLEGE OF CLINICAL PHARMACY 2021;5:554–69.

37. Smith ZR, Palm NM, Smith SE, et al. Critical care pharmacist perspectives on optimal practice models and prioritization of professional activities: A cross-sectional survey. Am J Health Syst Pharm 2024.

38. Sikora A MB, Most A, Martin G. Critical Care Pharmacists Save Lives. ICU Management and Practice 2024.

39. Sikora A. Critical Care Pharmacists: A Focus on Horizons. Crit Care Clin 2023;3:503–27.

40. Murray B, Newsome AS. Avoiding cost avoidance. Am J Health Syst Pharm 2022;2:14–15.

